# Monitoring and predicting SARS-CoV-2 epidemic in France after deconfinement using a multiscale and age-dependent model

**DOI:** 10.1101/2020.05.15.20099465

**Authors:** Jürgen Reingruber, Andrea Papale, David Holcman

## Abstract

The world and France were strongly impacted by the SARS-COV-2 epidemic. Finding appropriate measures that effectively contain the epidemic without putting severe pressure on social and economic life is a major challenge for modern predictive approaches. We developed an analytical framework to precisely monitor and predict the spread of the epidemic together with its impact on the health care system. The current implementation accounts for interactions between five age-stratified population groups, and predicts disease progression and hospitalization status using eight different categories such as infected, hospitalized, occupancy of intensive care units, deceased, recovered from hospitalization and more. We use a variety of public health care data for the five most infected regions of France during lockdown (March 18th till May 11th) to validate and calibrate the model. At day of deconfinement (May 11th), we find that around 14% (around 4.8M) of the population is infected in the five most affected regions of France (extrapolating to 5.8M for France). We then apply the calibrated model to explore different deconfinement scenarios. We find that wearing of masks and social distancing can prevent a significant second peak. In the context of school openings with limited testing capacities, we argue that testing should focus on children, but without tracing it will have only a limited impact. Finally, we explore a complementary deconfinement scenario where the fragile elderly population initially remains confined, while the rest of the population becomes gradually deconfined to achieve herd immunity within few month before deconfining also the elderly.

## 1 Introduction

The fast spreading worldwide pandemic of SARS-CoV-2 [1, 2, 3] has destabilized all major economies of the world in only a few months, forcing most European countries into confinement. This measure has curbed in few days the disease progression. To prepare the deconfinement and to recover economical prosperity, most of the affected western countries rely on the efficiency of wearing masks [4] and on social distancing, hoping that these measures will be sufficient to prevent a second peak of infection that could saturate hospitals and ICU. For such fast spreading and severe pandemic [5], predictive modeling is crucial to estimate in advance the impact of deconfinement measures, because small deviations can rapidly be exponentially amplified [6, 7, 8, 9, 10, 11]. Yet predicting with high accuracy remains a major challenge during this crisis [12, 13].

Very quickly after the beginning of the pandemic, public web sites have provided daily data for hospitalizations, ICU occupancy, deaths, recovered, etc.., which is crucial to obtain an age-stratified analysis [14]. The large number of severe cases in particular for the age groups 60-69 and older than 70 has destabilized the ICUs. But this data alone provides only a partial understanding of the mechanisms that govern the pandemic social propagation. It remains challenging to reconstruct the pandemic dynamics from this live data for several reasons: incomplete data, change in social interactions, or unknown fraction of the population that are asymptomatic, which are not accounted for in public health data.

In order to understand the dynamics of SARS-CoV-2 transmission and to predict the infection spread and the status of the health care system after deconfinement, we develop a discrete dynamical model that we implement for 5 different age groups (group 1=0-24, group 2=25-49, group 3=50-59, group 4=60-69, and group 5=70+) and 8 different infection categories (Tables S2 and S3). Because the distribution of infected in the 5 most affected regions (Île de France, Grand Est, Auvergne Rhône Alpes, Hauts-de-France and Provence-Alpes-Côte d’Azur) and the rest of France is very heterogeneous (the 5 regions together concentrate 36.9M people (67M for France) but account for 80% of the reported cases), we focused on these 5 regions to calibrate and validate the model, and to test several scenarios for the time after deconfinement. We found that the number of infected after lockdown is 14% (around 4.8M) in the five most affected regions and 5.8M of the total French population, in line with a recent report emphasizing the large number of asymptomatic in the population (15.5% from [15]) and slightly higher than a recent estimation of 3.7M infected [16]. Because of this low fraction of infected, a second catastrophic peak after deconfinement is unavoidable without proper control and social distancing measures. We found that wearing masks for the entire population could be a means to steadily prevent a second peak without the need of going through several re- and deconfinement phases. Moreover, we show that school opening poses a serious risk that could destabilize the deconfinement phase if social distancing measures and wearing of masks are not rigorously followed. If testing capacities are limited, we propose to focus testing on children to timely unravel the asymptomatic infected. However, we also find that testing without tracing only has a very limited effect on containing the pandemic. Finally, based on the analysis of the hospitalization data, we propose an alternative deconfinement scenario with the aim to quickly establish a protective herd immunity.

## 2 Results

The distribution of hospitalizations from public data reveals a strong heterogeneity of the infection spread throughout France (Fig. 1A-B). We therefore focused our analysis on data from the 5 most affected regions, which represent around 54% of the French population (36.7M) and around 80% of the reported cases. The age resolved time series for the number of hospitalizations, ICU occupancy, cumulative returned from hospital and cumulative deceased are very similar between the 5 regions and the entire country (Fig. 1D-E), and the ratio of these numbers is almost constant (Fig. 1F). Thus, our results and predictions for the 5 regions can be extrapolated to entire France by multiplying with a factor around 1.2 instead of a factor around 2 corresponding to the population ratio.

**Figure 1:**
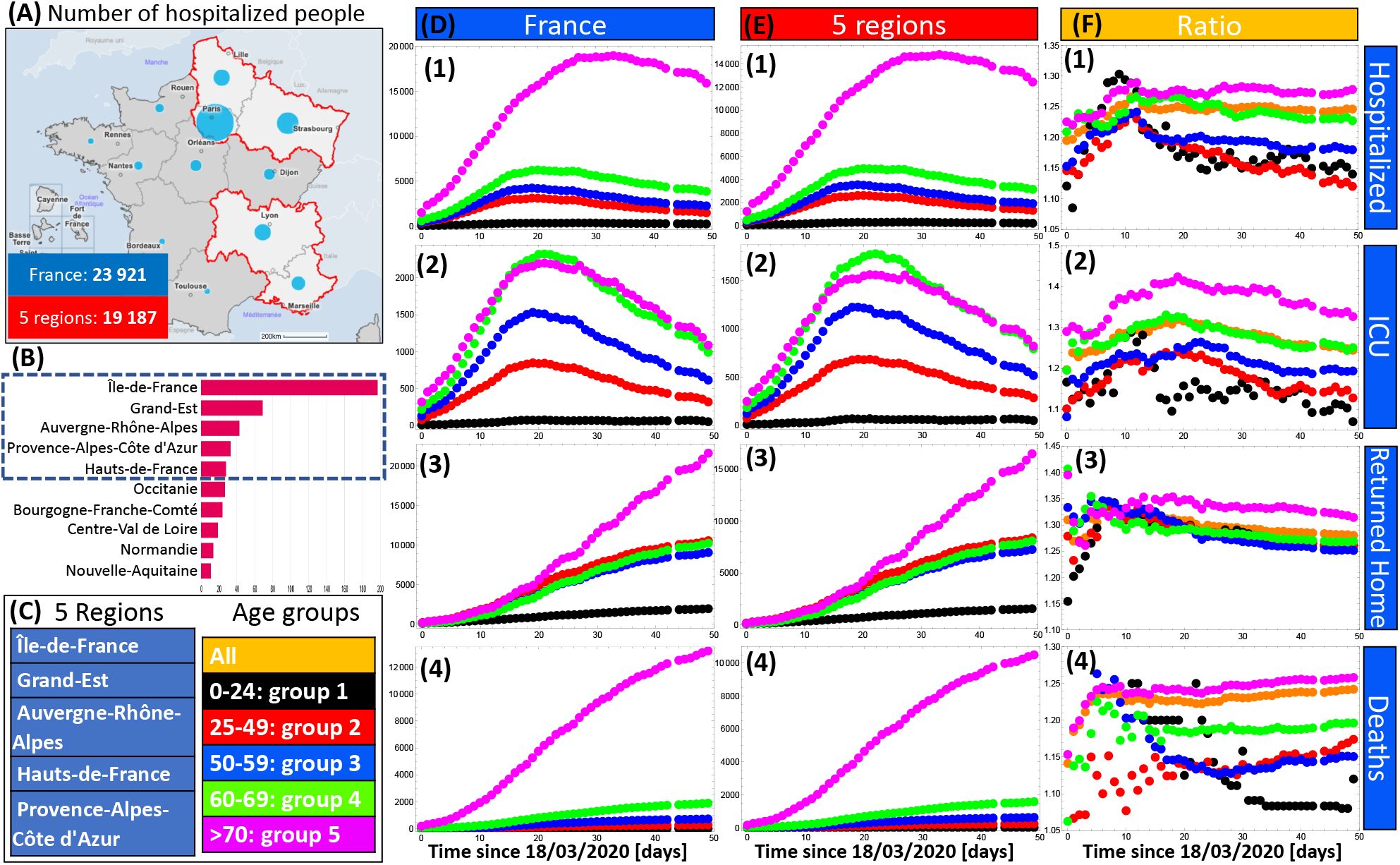
Public French health care data. (A) Map of France emphasizing the 5 most infected regions at 6th of May: Île de France, Grand Est, Auvergne Rhône Alpes, Hauts-de-France and Provence-Alpes-Côte d’Azur. (B) Distribution of people in ICU. (C) 5 Regions and age group stratification. (D-E) Age resolved time series for hospitalized (1), ICU occupancy (2), cumulated recovered from hospitalization (3) and cumulated deaths (4) for France and the 5 regions. (F) Ratio between the data in D (entire France) and E (5 regions). The color code for the age groups is defined in panel C. Data source [17, 18].

### 2.1 Model calibration with public data

Our discrete and spatially homogeneous transition model has a time resolution of one day and accounts for three different time scales to compute the disease progression: the normal time, the time since infection and the time since a person switched to its current infection category (Fig. 2A). To account for age dependent characteristics, we partitioned the population into 5 different age groups (group 1=0-24, group 2=25-49, group 3=50-59, group 4=60-69, and group 5=70+). To model the disease progression we consider 8 infection categories (Fig. 2B-C, see also Tables S3 and S2 in the SI) and we use transition probabilities to compute the dynamics of the disease (Fig. 2C). All parameters can be updated at any day to account for modified conditions (e.g. confinement or hospitalisation procedures). For more details we refer the reader to the Methods section in the SI.

**Figure 2:**
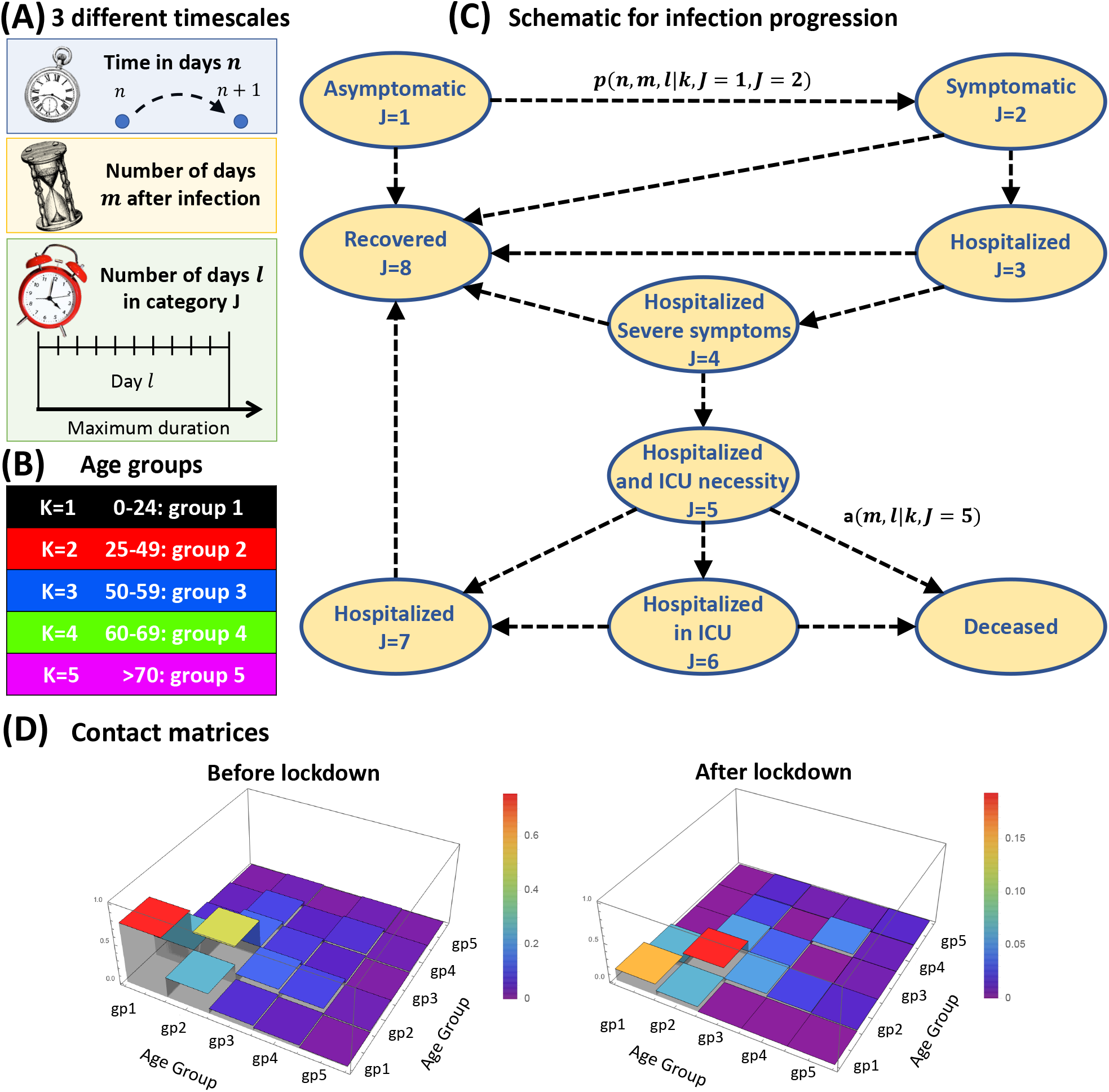
Model description. (A) The time resolution of the discrete model is one day. The dynamics depends on three timescales: the normal time (*n* in days), the time since infection (*m*) and the time since an infected joined its current infection category (*l*). (B) Definition of the 5 age groups considered in the model. (C) Model for disease progression with transitions between the 8 infection categories. The parameters *p*(*n, m, l*|*k, J_old_, J_new_*) are the multi-timescale transition probabilities to switch from category *J_old_* to *J_new_*. The parameters *a*(*m, l*|*k, J*) are the decease probabilities. Infected persons only die in the hospital in categories *J =* 5 (waiting for ICU) and *J =* 6 (in ICU). (D) Normalized contact matrices before and after lockdown. For more details please consult the Methods section in the SI.

To model the social interactions before lockdown, we use the contact matrix between age groups in France from [19] ((Fig. 2D). Since before lockdown we did not have access to age stratified data, we only calibrated the exponential growth rate for the number of new infections (0.18*day*^−1^, see Fig. S1) by modifying the infectiousness parameter *β* (see Eq. 6 in the Methods; we assume that the infectiousness is independent of age [7]). Using *β* and the contact matrix from [19] we found for the time before confinement for the five groups the reproduction numbers [3.8, 3.1, 2.7, 2.4, 0.5] (see Eq. 7 in the Methods). The reproduction numbers are different for each group because of the internal structure of the contact matrix. For the averaged collective reproduction number, we obtain 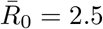 (see Eq. 8). If we account only for the young and active population (groups 1-3) that make most of the social contacts, we get 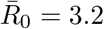, similar to [16, 20]. To conclude, to accurately monitor the pandemic one has to consider age dependent reproduction numbers.

After lockdown, we used the age-stratified public data for the 5 regions [17, 18] for model calibration. We first modified the contact matrix to account for the reduction in social interactions (see Methods and Fig. 2C). We then tuned our model parameters (e.g. probability to become hospitalized, probability to develop severe symptoms, decease probabilities etc) to obtain simultaneous agreement with all the available data (Fig. 3). Interestingly, the very different behaviour of the hospitalization data for group 5 could only be accounted for by gradually increasing the duration of hospitalization during lockdown starting around 10 days after lockdown, which probably reflects change in the policy of hospitalization for this group (personal communication). Although most of the model calibration for group 5 matches well with the data (magenta curves in Fig. 3), the model still overestimates the cumulated number of recovered for this group (Fig. 3 KJ) starting around day 20 after confinement. The exact reason is unclear. A possible explanation could be that the contact matrix during lockdown is not constant but the number of interactions especially for group 5 further decreased during the lockdown, and without this adaptation we overestimate the hospitalization influx and outflux for this group. At the end of the lockdown period, the model predicts that around 14% of the population has been infected (Fig. 4A), most of them in group 1 and 2 (Fig. 4B). A 5% fluctuation in 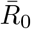 leads to a variation in the fraction of infected at the end of the lockdown in the range of [11% – 17%] (Fig. S2A-C). Finally, due to the reduced social interactions, the reproduction numbers for the five groups during lockdown are reduced to [0.73, 1.08, 0.74, 0.67, 0.25]. The collective reproduction number is 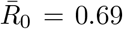, which is around a factor of 4 smaller compared to pre-lockdown, in agreement with [16]. To conclude, the calibrated and validated model faithfully reproduces the all the available data, and we use it now to study the pandemic progression and its load on the health care system for the deconfinement period after lockdown.

**Figure 3:**
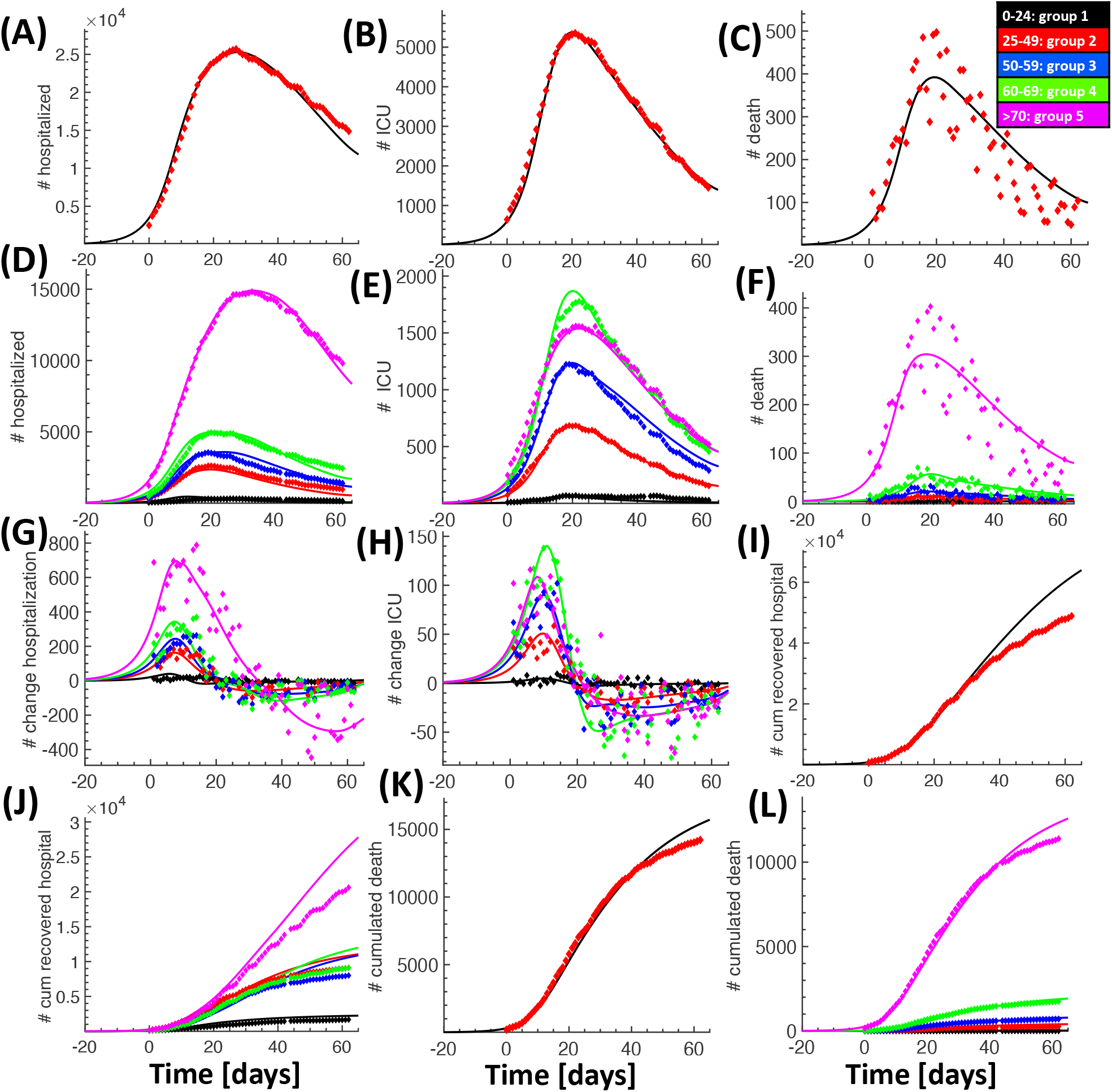
Calibration of the model using age-stratified data for the 5 regions after lock-down. We compare data (diamonds) with simulation results (continuous lines). Number of hospitalized (A), ICU occupancy (B), daily deaths (C), and their age group distributions (respectively D, E and F); daily change in hospitalization (G) and ICU (H) per age group; cumulative number of people recovered from hospitalization (I) with age distribution (J), and cumulative number of deaths in hospitals (J) with age distribution (L). Day zero corresponds to the 18th of March.

**Figure 4:**
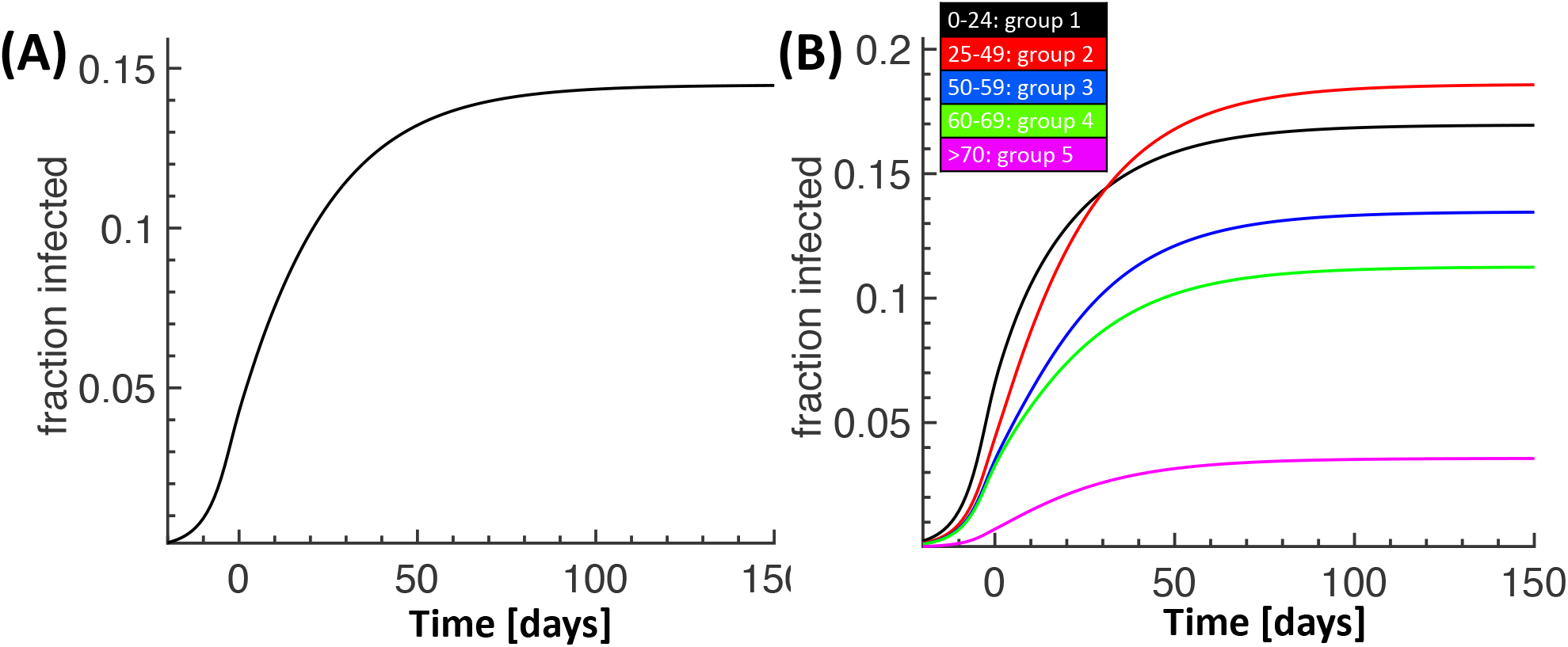
Fraction of infected without deconfinement. Predictions of the fraction of infected population for the 5 regions (A) and its age group distribution (B). Day zero is March 18th.

### 2.2 Scenario for the pandemic evolution without social interventions

We used the calibrated model to show the drastic consequences for the hypothetical case that no social measures would have been taken to contain the pandemic (Fig. 5). In this case we predict a total of 250,000 deaths in the 5 regions (Fig. 5D), which would extrapolate to ~450,000 deaths in France. In this uncontrolled case the majority of the population would become infected (around 87%, Fig. 5A), and the ICU would remain saturated for around 50 days (Fig. 5C). Age stratified simulation results for this scenario are shown in Fig. S3.

**Figure 5:**
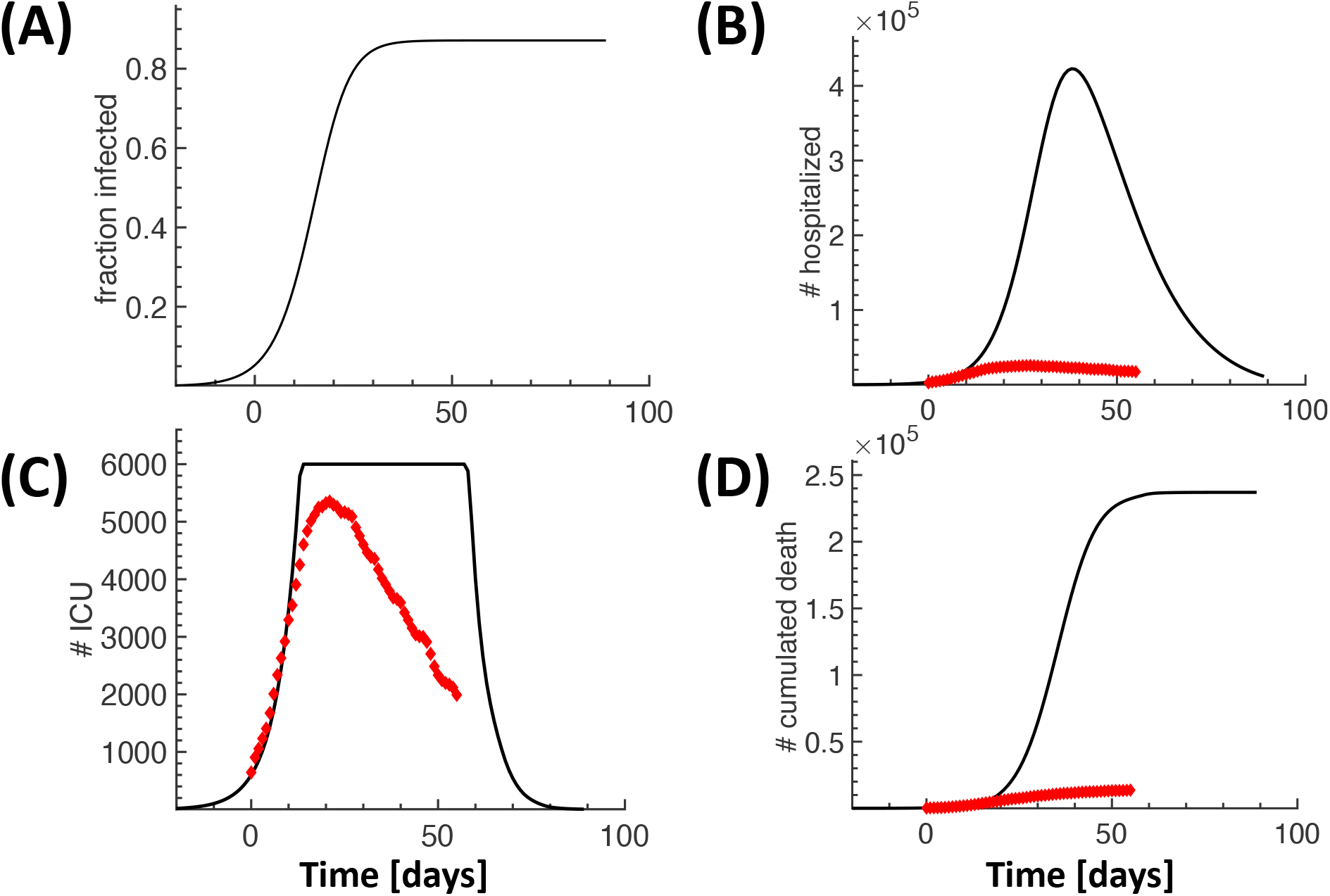
Pandemic spread in absence of social interventions and restrictions. Simulation results that show the drastic consequences for the hypothetical scenario that no social measures would have been taken to contain the spread of the pandemic. Continuous lines show simulation results, the red dots are the data from Fig. 3. New infections (A), hospitalizations (B), ICU occupancy (C) and cumulative deaths (D). Day zero is March 18th. We used maximal number of 6000 ICU beds in the five regions.

### 2.3 Deconfinement scenario after lockdown in absence of social restrictions

If all social restrictions would be alleviated after the lockdown and social interactions would return to the same level as before confinement, we predict a large second peak for the new infections (around 6 times the first one) that would occur around mid-July (Fig. 6A) with around 220,000 cumulated deaths for the 5 regions at the end (Fig. 6C). ICU would be saturated for 50 days (Fig. 6B), with a need 320,000 hospitalizations at the peak (Fig. S4B).

**Figure 6:**
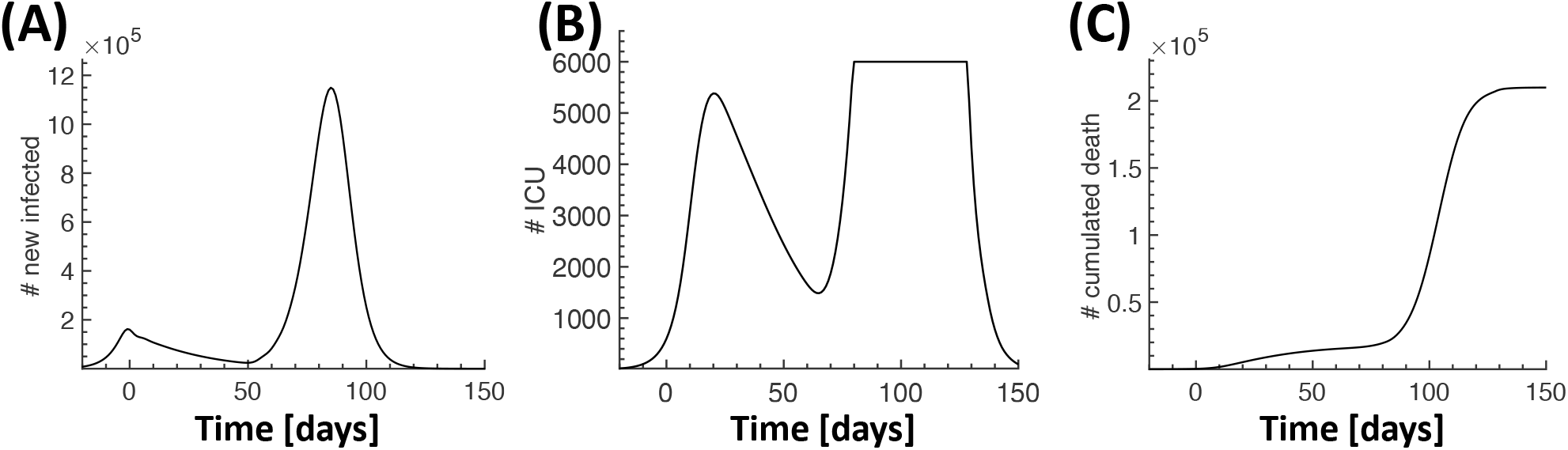
Deconfinement scenario without social restrictions. Simulation results for the case that social life after deconfinement would be the same as before confinement. New infections (A), ICU occupancy (B) and cumulative deaths (C). Day zero is March 18th.

### 2.4 Controlling the pandemic after deconfinement by wearing masks

Several evidences suggest that wearing a mask could efficiently attenuate the epidemic spread [4] by reducing the propagation of viral particles due to breathing, coughing or sneezing, as revealed by a recent analysis using Laser Light Scattering [21]. However, as demonstrated in the case of influenza virus, coarse and fine droplets with diameters > 5*μm* and < 5*μm* are not reduced by the same proportion by wearing masks. Whereas course droplets are reduced by a factor of 25, fine droplets are reduced only by a factor of 2.8 [22]. Since fine droplets contain around ten times more viral particles than course droplets, and they stay much longer in the respiratory environment [22], we hypothesize that wearing masks could reduce the infectiousness by a similar factor. We thus explored how the pandemic would spread if the social interactions after deconfinement would be the same as before lockdown, but wearing of masks would reduce the infectiousness *β*, and thus the reproduction number 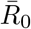. In Fig. 7 we tested three scenario where 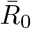 is reduced by a factor of 3 (strict mask wearing, red curves), 2.5 (less strict, blue curves) and 2 (insufficient, green curves). For comparison, we also show the results for the no-deconfinement scenario (black curves) and the scenario with full deconfinement (magenta curves). The cumulated number of deaths for the 5 regions is 21,000 (*R*_0_*/*3), 35,000 (*R*_0_/2.5 and 70,000(*R*_0_/2) (Fig. 7F) with 16%, 30% and 50%) of the population infected (Fig. 7A).

**Figure 7:**
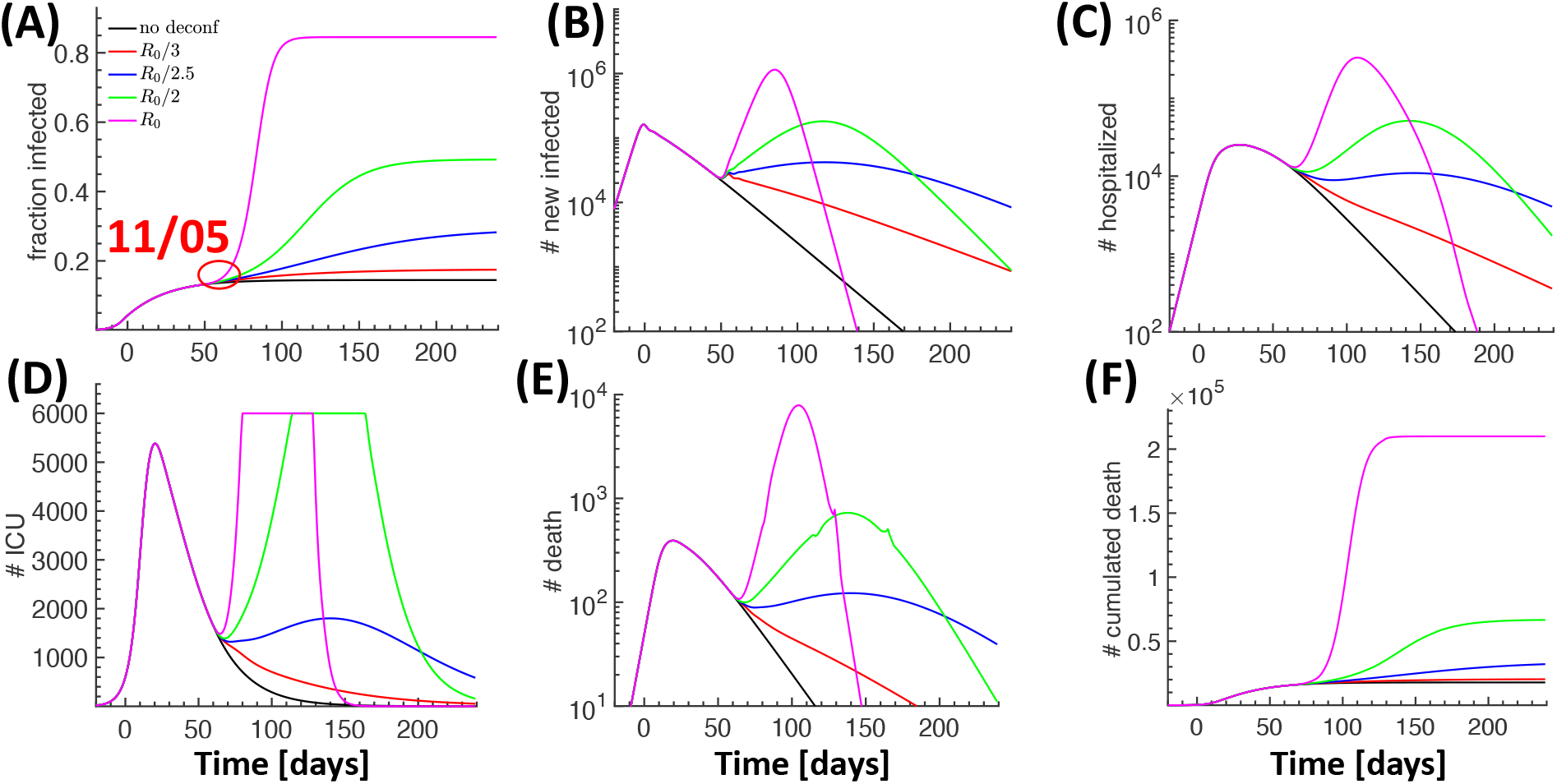
Effect of wearing masks after deconfinement. Wearing masks corresponds to a reduction in infectiousness represented by the reproduction number *R*_0_. Comparison of three scenarios where *R*_0_ is reduced by 3 (strict mask wearing, red curves), 2.5 (less strict, blue curves) and 2 (insufficient, green curves). The results for no deconfinement (black curves) and deconfinement without social restrictions and mask wearing (magenta curves, no reduction in *R*_0_) are also shown for comparison. Fraction of infected people (A), daily new infected (B), hospitalized people (C), ICU occupancy (D), daily deaths (E), and cumulative number of deceased (F). Day zero is March 18th.

Next we explored the consequences that children under 11 are not obliged to wear masks at school. We assume that adults (group 2 -5) strictly wear their masks resulting in a reduction of their infectiousness by a factor of 3, whereas the reduction for group 1 is only by a factor of 2 (Fig. 8, red curves) or 1.5 (Fig. 8, blue curves). We found that with a reduction by a factor of 2 the deconfinement phase remains under control with no large second peak and ICUs remain unsaturated (Fig. 8A-D, red curves). However, by only slightly increasing *R*_0_ for group 1 by a factor of 1.33 a large second peak emerges (Fig. 8A-D, blue curves). The number of hospitalized would reach 540,000 beginning of August, ICU would start to be saturated around beginning of July and the cumulated number of deceased would reach around 60,000 (around 3 times the level of end of April in the 5 regions). This suggests that the behaviour of group1 could destabilize the deconfinement phase if not carefully controlled.

**Figure 8:**
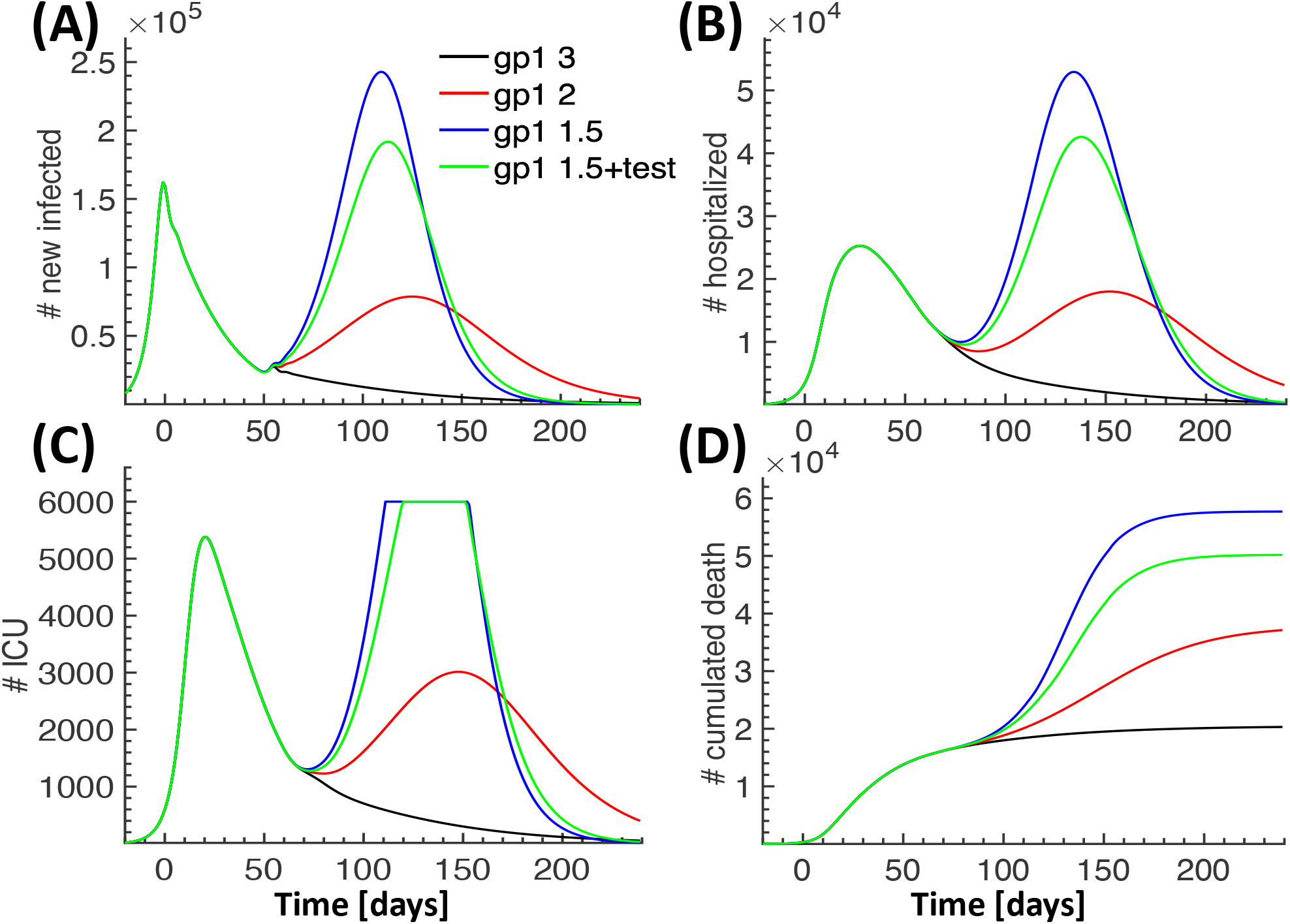
Effect of a reduced susceptibility for mask wearing in group 1. Comparison of four scenarios for the time after deconfinement where the infectiousness for group 2-4 is reduced by a factor of 3, and for group 1 it is reduced by a factor of 3 (black curves, corresponding to the red curves in Fig. 7), 2 (red curves) and 1.5 (blue curves). In the forth scenario (green curves) we further explored the impact of testing for group 1. New daily infected (A), hospitalized (B), ICU occupancy (C), cumulative number of deaths (D). Day zero is March 18th.

Finally, we explored whether testing could reduce the second peak for the case that the infectiousness of group2 is only reduced by a factor of 1.5 (Fig. 8A-D, green curves). We assume that 100,000 tests can be made every day, and an infected person that has been tested it is removed from the pool of infectious the day after testing. Because the infectious population in group 1 generates the second peak, simulations (not shown) reveal that focusing all the testing capacities on group 1 is most effective. However, even in this case (green curves in Fig. 8) the second peak would only be slightly reduced and the health care system would still be destabilized with 50,000 deceased people after 150 days post confinement (mid-August).

We conclude that whereas wearing masks is efficient to keep the pandemic under control, testing without tracing has only a limited impact. In addition, it is problematic to trace children that usually have no smartphones. Thus, school openings without efficient control can destabilize the deconfinement phase.

### 2.5 Deconfinement scenario with controlled gradual restoration of social interactions and careful protection of elderly

Because the number of hospitalized, present in ICU and deceased is highest for elderly people belonging to groups 4 and 5 (Fig. 3), a major challenge is to protect them during the deconfinement phase. We now present a deconfinement scenario occurring in three phases (Fig. 9): During phase 1 (lasting 75 days) groups 1-3 are fully deconfined and their infectiousness is reduced by a factor of 2 for group 2-3 and by 1.5 for group 1 due to mask wearing. Groups 4-5 remain strictly confined and protected to avoid infections. In phase 2 (lasting 70 days) all restrictions for groups 1-3 are removed and they are no longer required to wear masks. Groups 4-5 still remain strictly confined. Finally, in phase 3 groups 4-5 are fully deconfined and all social life returns to the level before lockdown.

**Figure 9:**
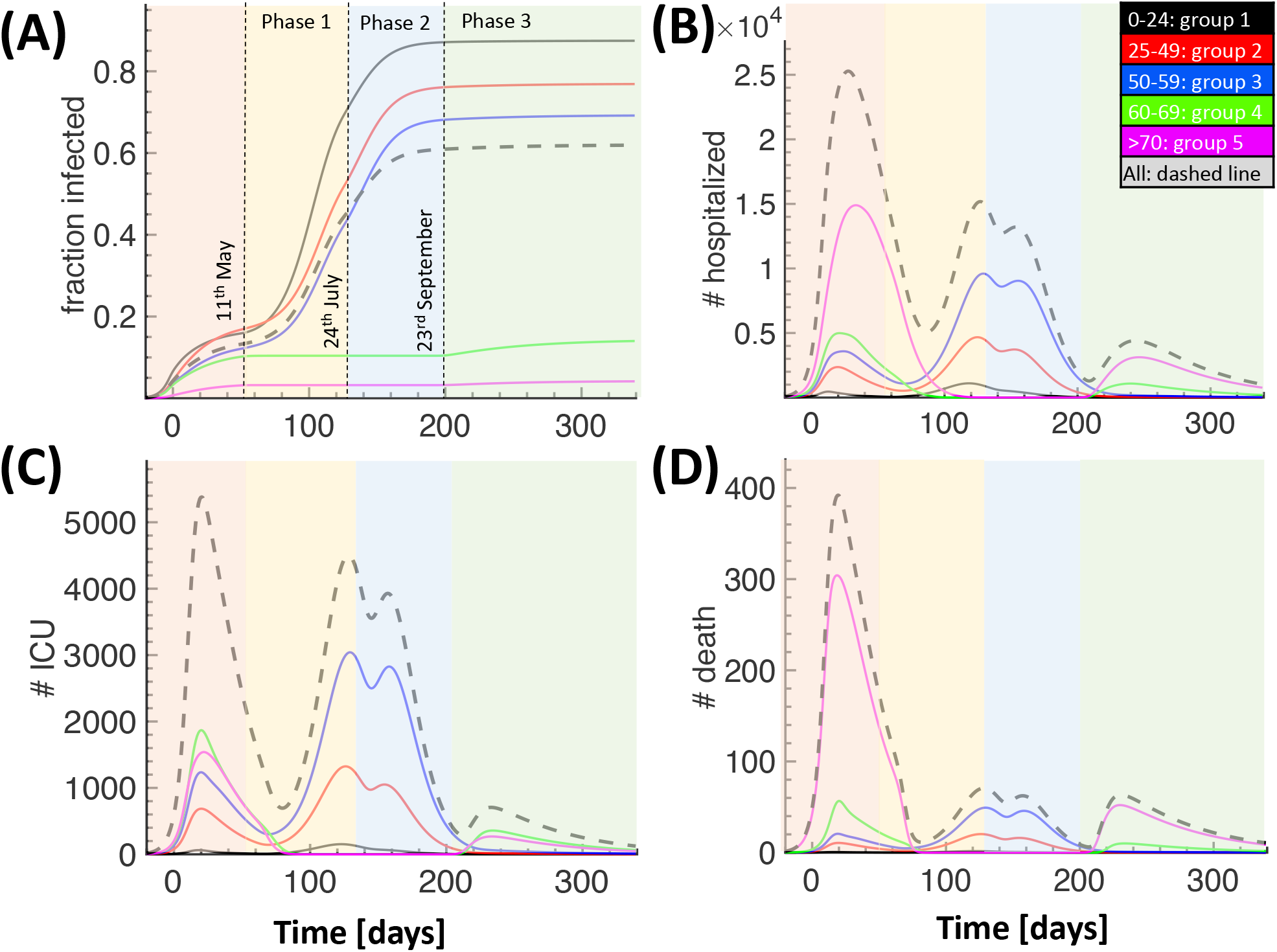
Deconfinement scenario with careful protection of elderly and controlled gradual restoration of social interactions in three phases within less than half a year. In phase 1, groups 1-3 are fully deconfined and their infectiousness is reduced by a factor of 2 for group 2-3 and 1.5 for group 1 due to mask wearing. In phase 2, groups 1-3 are no longer required to wear masks. Groups 4-5 stay strictly confined and protected during phase 1-2 to avoid infections. Finally, in phase 3 also groups 4-5 are fully deconfined and all social life returns to the level before lockdown. For more information see text. Fraction infected per age group (A), hospitalized (B), ICU occupancy (C) and number of deceased (D). Day zero is March 18th.

During phase 1, the contact matrix for group 1-3 is the same as before lockdown, but we consider that wearing masks moderately reduces the infectiousness (respectively *R*_0_) of group 2-3 by a factor of 2, and for group 1 by 1.5. During phase 1 the number of hospitalized (Fig. 9B) increases again, however, the ICU remain unsaturated (Fig. 9C). Although the number of death increases slightly, it remains at a low level due to the low probability to die for groups 1-3 (Fig. 9D). Most important, during phase 1 the fraction of infected increases to a level that gives herd immunity (Fig. 9A). At the end of phase 1 the number of people hospitalized and in ICU decreases again, and this is when phase 2 is initiated where we additionally remove the reduction in infectiousness for groups 1-3 (no necessity to wear mask). For these groups the situation in phase 2 is therefore identical to the one before lockdown. In contrast, groups 4-5 remain strictly protected. Although in phase 2 the simulations show an initial increase in hospitalized, ICU and deceased, there is no exponential growth due to the high level of herd immunity, and the numbers quickly decline again (Fig. 9B-D). The fraction of infected in groups 1-3 further increases during phase 2, thereby strengthening the herd immunity. Finally, when phase 3 starts groups 4-5 are fully deconfined. The social interactions between all groups are now the same as before lockdown. This deconfinement of groups 4-5 initially slightly increases the number of hospitalized, ICU and death, however, due to the large immunisation in the other groups there is no danger of exponential growth and the situation quickly normalizes. To reduce the number of hospitalized and death, the infectiousness during phase 1 could be further reduced by imposing stricter rules for mask wearing, for example. However, a drawback is that these measures would prolong the overall time until sufficient herd immunity is reached such that group 4-5 can be deconfined. To conclude, this deconfinement scenario in three phases would quickly achieve herd immunity and return to normal social and economic life with a low number of death and a manageable load on the health care system.

## 3 Discussion

Preventing the natural exponential spread of the COVID-19 pandemic is a major challenge of the deconfinement phase. Using public health care data, we developed a novel modeling approach that combines social interaction matrices with dynamical modeling to reproduce and predict the time course of the pandemic together with its detailed consequences for the health care system. A major strength of our approach is to simultaneously account for a large variety of age-stratified data such as hospitalization, ICU occupancy, recovered from hospitalization, which provides strong confidence for model predictions.

In most European countries and France, lockdown has reversed the exponential spread of the pandemic into a decline. However, since the fraction of susceptible after lockdown is still very large, and asymptomatic persons are difficult to reveal, it remains a major challenge to avoid a return to an exponential growth phase with similar rates as before lockdown. Our dynamical model allows to explore in detail the time evolution of the pandemic for various scenarios, and thus provides a tool to refine policies. By continuously adapting the model parameters to real time data, simulations can be used to predict the evolution of the pandemic in order to test and implement in advance efficient social measures to control and contain the pandemic spread.

In the absence of social measures a catastrophic second wave is unavoidable [16, 20, 8], however, we found that wearing masks for the entire population could prevent this to happen without the need to go through several re- and deconfinement phases. In that context, we find that school openings without rigorous control of mask wearing and social distancing poses a serious risk to destabilize the deconfinement phase. If testing capacities are limited, we further argue that the focus should be on testing school children to timely unravel the asymptomatic infected. A drawback of wearing masks is that the pool of susceptible decreases only slowly, and therefore the threat of a second wave persists. Since vaccines will probably not be available before end of the year, this requires to carefully monitor the situation for a long time. Based on the analysis of the hospitalization data, we propose an alternative deconfinement scenario with the aim to quickly establish a protective herd immunity: in the first part lasting until end of September, only the population below the age of 60 will be gradually fully deconfined with the need of mask wearing until end of July, and by relieving all restrictions afterwards. Finally, when the elderly population above the age of 60 is also deconfined end of September, this poses only a minor and controllable risk because of the large immunisation in the rest of the population.

## Data Availability

public data available online (see url below)

https://www.data.gouv.fr/fr/datasets/donnees-hospitalieres-relatives-a-lepidemie-de-covid-19/

https://geodes.santepubliquefrance.fr/\#c=news

## Supplementary Information

### Methods

#### Model description

We developed a discrete and spatially homogeneous mean-field transition model with a single day as time-resolution. Although we implemented the model for 5 age groups and 7 infection categories (Tables S2 and S3), the model structure is general and can integrate a larger diversification such as refining the age groups and infections categories, or distinction between female and male.

The present approach differs from SIR, SEIR or associated models, as it incorporates three different time scales: the usual day counting (labeled by *n*), the time since infection (*m* days) and the time since an infected joined its current infection category (*l* days). For example, *m* = 1 specifies that a person has been infected yesterday, and an infected that is in ICU (*j* = 6) since 10 days has *l* = 10. Whenever a person switches to a new category, the timer *l* is reset to one, otherwise it is increased. With these three time clocks we can incorporate very heterogenous hospitalization procedures such that we re able to precisely monitor and predict the evolution of hospital and ICU occupancy. We can model heterogeneous hospitalization conditions without the need to alter the model structure by adding new variables. For example, we can incorporate the distribution of time spend in ICU by using a multi-modal probability distribution that depends on *l*, without introducing different ICU populations. The model structure also allows to implement correlations between the three time clocks *n*, *m* and *l*. For example, the fate of a person that is in ICU since *l* days may as well depend on the time *m* after the infection. In principle, all model parameters can be functions of n, m and l.

#### Variables

- *S*(*n*|*k*): Number of susceptible persons at day n belonging to group k.
- *I*(*n, m,l*|*k, j*): Number of infected persons belonging to group k, infection category j, day m after infection and day *l* in infection category j.
- *I_inf_* (*n, m, l*|*k, j*) *= ξ*(*m, l*|*k, j*)*I* (*n, m, l*|*k, j*): Infectious persons selected from the pool of infected. We consider here that only asymptomatic persons (category *j* = 1) are infectious between *m* = 5 − 11 days after infection (5 days of incubation time) (see Table S6). We are aware that it is difficult to give a precise definition of what exactly are asymptomatic persons. We assume that asymptomatic persons that switch to category *j* = 2 (with symptoms) have such kind of symptoms that they take measures to quarantine themselves to avoid infecting others. With this definition, paucisymptomatic persons that are not aware of their infection, or show only mild symptoms such that they do not take measures to quarantine themselves, would still belong to the asymptomatic category. However, a different selection matrix *ξ*(*m*, *l*|*k, j*) could be implemented such that also symptomatic persons in category *j* = 2 are infectious during some initial days after they start showing symptoms. We assume infected from group 1-3 and group 4-5 start to show symptoms between days *m* = 6 − 10 with overall probabilities 0.8 and 0.9, respectively (see Table S4).
- *I_new_*(*n*|*k*): Number of new infected at day n belonging to group *k*. The next day these persons are added to the pool of asymptomatic infected, *I*(*n* + 1,1,1|*k*, 1) = *I_new_*(*n*|*k*)
- *D*(*n, m,l*|*k, j*) = *a*(*m,l*|*k, j*)*I*(*n, m, l*|*k, j*): Number of deceased belonging to group *k* that die in category *j* at day *n*, *m* days after the beginning of their infection, and *l* days in category *j*. *a*(*m,l*|*k*, *j*) is the decease probability.
- *R*(*n, m, l*|*k, j*) = *p*(*n, m, l*|*k, j, J_max_*)(*I*(*n, m, l\k, j*) − *D*(*n, m, l*|*k, j*)): Number of persons belonging to group *k* that recover from category *j* at day *n*, *m* days after infection, and *l* days spent in category *j* before recovering.

The algorithm for the time evolution with ∑(*n, m, l*|*k, j*) *= I*(*n, m, l*|*k, j*) *− D*(*n, m, l*|*k, j*) is

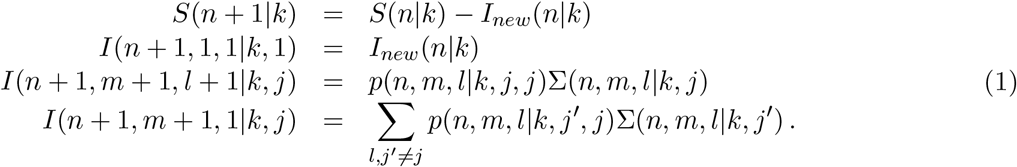

The transition probabilities *p*(*n,m,l*|*k,J′, J*) satisfy the normalization condition 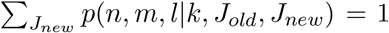. With the cumulative number of dead and recovered persons per group

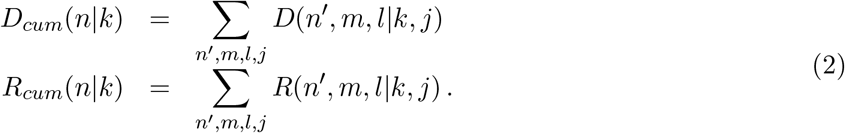

we obtain the conservation equation

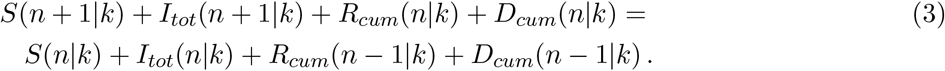

#### New infections

To estimate the number of new infections, we consider the number of contacts *c*(*n*|*k, k′*) that group *k* and *k′* make at day *n*. The total number of contacts group *k* make is *c*(*n*|*k*) = ∑*_k′_ c*(*n*|*k, k′*). The number of infectious persons in group *k* is

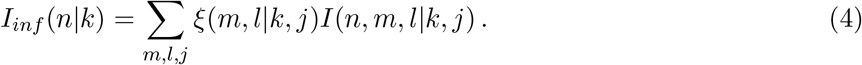

The amount of new infections in group *k* generated by the infectious in group *k′* is

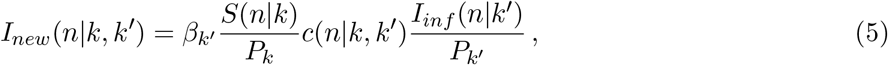

where *P_k_* is the population in group *k* and *β_k_* the infectiousness of group *k*. The total number of new infected in group *k* is

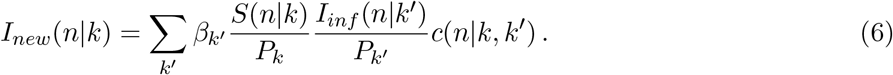

The parameter *β_k_* describes the infectiousness per group. *β_k_* can be time dependent, for example to account for an increasing fraction of people that wear masks. Because there is no solid evidence that infectiousness depends on age [7], we assume for the time before and during lockdown (where masks were largely unavailable) that *β_k_* = *β* is constant and independent of age. After lockdown, *β_k_* changes due to mask wearing and can be group dependent, e.g. because the fraction of people that wear masks might differ between the groups.

#### Reproduction numbers

In this paragraph we discuss some properties of the group dependent reproduction number in relation to the parameter *β* described above. With group independent *β_k_* = *β*, to connect *β* to reproduction numbers we consider the initial condition with 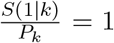 and a single infectious person in group *i*, *I_inf_*(1|*k*) = *δ_k,i_*. The total number of infected generated by this person per day is

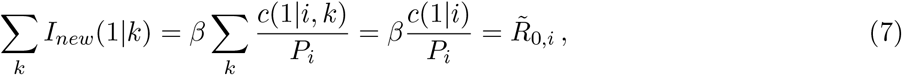

where 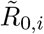 is the reproduction number per day. Eq. 7 shows that if the number of contacts is proportional to the group size, 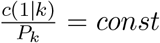, then 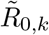 is the same for all groups. Moreover, Eq. 7 together with Eq. 5 reveal that the number of new infected is independent of the exact normalization of the contact matrix.

To connect the daily 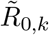 to the classical reproduction number 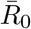, we consider that an infected person is infectious for in average 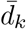 days. The mean reproduction number per infections period is then 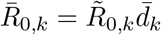, and the averaged collective reproduction number is

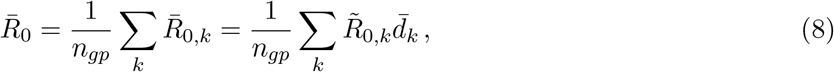

where *n_gp_* is the number of groups.

In our implementation we assume that an infected person starts to develop symptoms between days 6 − 10 after infection (Table S4) with overall probability *P_k_* (*Q_k_* = 1 − *P_k_* is the probability of remaining asymptomatic), in which case he is put in quarantine the next day. With this information, the mean number of days an infected person remains infectious is

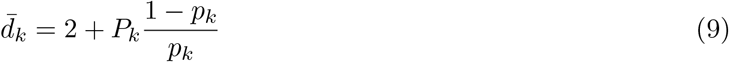

where 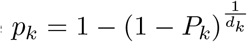 and *d_k_* = 5. The factor of 2 in Eq. 9 accounts for the fact that a person that starts to show symptoms is removed from the pool of infectious only the following day. With 20% asymptomatic for group 1-3 and 10% for group 4-5 (Table S4) we get *d_k_* = [4.1, 4.1, 4.1, 3.5, 3.5].

#### Contact matrices

The contact matrix before lockdown is derived from the frequency matrix in [19] (S3 Table *Base-case contact matrix with age categories for all contact and for skin contact only*) by summing over age-groups to comply with our age-group definitions. The resulting normalized contact matrix is

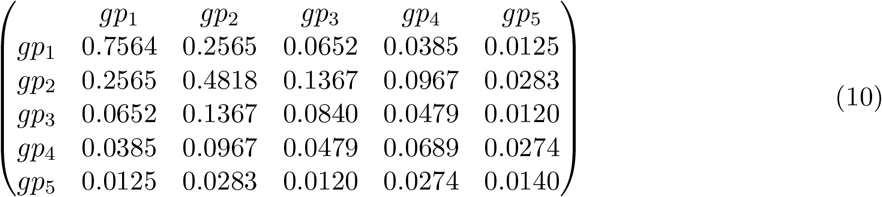

We chose group 2 as a reference group and normalized the contact matrix is such that the total number of contacts for group 2 is one, ∑*_k_ c*(2, *k*) = 1. To estimate the reduced contact matrix after lockdown we used the following reduction matrix *r*(*k*, *k*′):

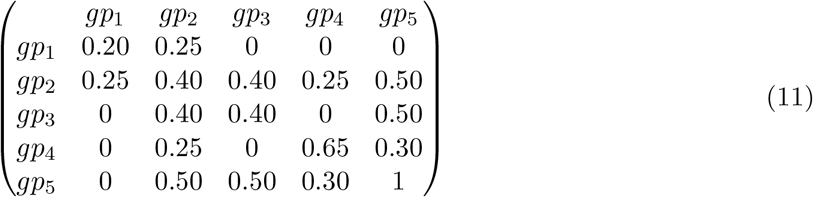

The resulting contact matrix *c_ld_*(*k*, *k*′) = *c*(*k*, *k*′)*r*(*k*, *k*′) after lock-down is:

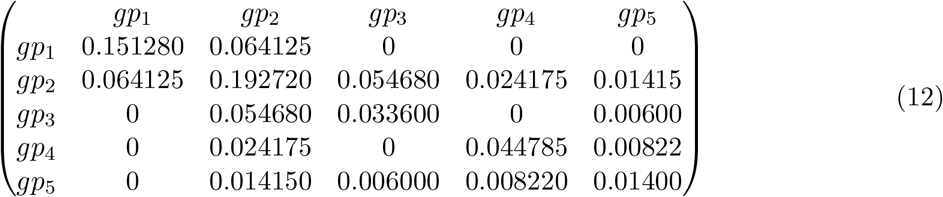

To build the reduction matrix, in the absence of available data, we made several ad-hoc assumptions motivated by the following arguments: the intra-group contacts for group 1 are mostly redundant and therefore strongly reduced by 80% due to the lockdon and closing of schools and universities. The inter-group contact between group 1 and group 2 is reduced to 25% of the original contacts to maintain mostly parents children interactions. In contrast, since the contacts made by group 5 are much less compared to the other groups, we assume that these contacts are mostly important and therefore cannot be as strongly reduced as for group 1 or 2. We assume no contacts between group 1 and 5 (complete disruption of the relation with grand-children), however, we kept 50%contacts with groups 2-3 due to social needs. Finally, we tuned values to obtain agreement with number of hospitalized and recovered during lockdown, which also leads to constraints. However, we hope that based on recent surveys we will soon have more precise estimations for contact matrices, which we will incorporate to refine the model.

#### Parameter estimations

A highly difficult task and major achievement was to identify a consistent parametrization that is compatible with all the available data related to the infection properties and the health care status. The parameter values used for the simulations are summarized in Tables S4, S5, S6. Initially we extracted published parameter values from [20, 16, 8], which we adjusted using the age-stratified data.

To model the social interactions before lockdown we use the contact matrix Eq 10. Since before lockdown we did not have age stratified data, we only estimated the value of *β* to obtain the exponential growth rate for the number of new infections (0.18*day^−^*^1^, see Fig. S1). However, instead of varying *β* itself, we chose group 2 as a reference and varied the daily reproduction number for group 2 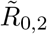. With Eq 7 we then have 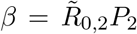 (note that we chose the normalization of the contact matrix such that *c*(1|2) = ∑*_k_ c*(1|2, *k*) = 1). By fitting the exponential growth we estimated 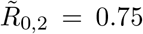. The daily reproduction numbers for all the groups are 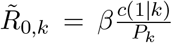, which gives (0.93, 0.75, 0.66, 0.69, 0.15).

After lockdown we used the age-stratified public data to adjust the transition and decease probabilities in order to simultaneously obtain good agreement with all the data, as shown in Fig. 3. To reduce the number of free parameters, we used time periods where switching between categories occurs together with overall transition probabilities for these periods. Assuming that transitions occur uniformly distributed, during these transition periods, we computed the daily transition probabilities that we then used for the simulations (Tables S4).

Many parameters are correlated leading to algebraic constraints: for example, a reduction in infectiousness can be compensated by an increase in the probability of hospitalisation, such that the number of hospitalized remaind unchanged. Such ambiguity could be resolved by using precise data (still unavailable) for the contact matrices and the virulency. Nevertheless, it is extremely difficult to identify a correct set of parameters that consistently reproduces all the age-stratified data that we used to calibrate and constrain the model (Fig. 3). These severe constraints gave us confidence that most of our parameters are reliable, simply because they faithfully reproduce large amount of diverse data. We are also aware that there are parameters that are not at all constrained by the current data and therefore their values in Table S4 may be taken with caution. For example, the hospitalization data does not constrain the time after which an asymptomatic person can be considered as recovered, and our choice for the time windows to switch from *J* = 1, 2 to *J* = 8 in Table S4 are largely arbitrary. But on the other hand these parameters do not affect the hospitalization predictions, which is our focus. Such parameters might be estimated in the future using new data, e.g. serological results. Moreover, it is very difficult to precisely estimate transition periods, here more detailed hospitalization data will be necessary to derive a more a precise model. Transition periods are less important for the number of death, but they are important to precisely estimate the dynamics for the occupancy of ICU and hospitals.

#### Testing

Since in our model only asymptomatic persons infect others, we use testing to reveal these persons. Given the available testing capacity per day, and assuming that testing occurs randomly among the population background consisting of susceptible and asymptomatic persons, we calculate the probability that asymptomatic infected are revealed by testing. These tested persons are then removed from the pool of infectious the next day.

#### Sensitivity analysis

Due to emergency of the COVID-19 crisis and the large dimensional parameter space, we performed a first sensitivity analysis only for *R*_0_ and the initial number of infected people (Fig. S2). We first vary evaluated the consequences of ±5% fluctuations of the parameter *β* (corresponding to varying the reproduction number 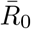 by the same fraction) for a scenario with lockdown and no deconfinement (Fig. S2A-C). In these three simulations, we used exactly the same parameters, except for *β*. Moreover, to correctly compare the consequences of changing *β*, in all simulations, the lockdown is initiated when the number of deceased patients reaches the same threshold. Because a larger *β* leads to faster exponential growth of infected and deceased, this threshold is reached faster if *β* is larger. Therefore the day when the lockdown is initiated has to be adjusted with *β*: it is 4 days earlier for the larger value of *β* compared to the lower value. We further aligned the simulation time such that the lockdown corresponds to the same calendar date. We find that varying *β* by 5% leads to a variation in the fraction of infected by around 18% with respect to the original value around 14.4% (Fig. S2A). For the upper value of *β* the ICU occupancy would reach the limit of 6000 ICU beds (Fig. S2A), and the peak number of deaths would increase by around 11 % (Fig. S2C).

By varying the total number of initially number of infected people by 20%, we use the same procedure as described above. Interestingly, the dynamics of fraction of infected, in ICU and deceased is quite insensitive to this perturbation, as the various curves are almost identical (Fig. S2D-F). At this stage, we conclude that changing the initial conditions only shifts the simulation time (2 days difference between upper and lower condition) when the lockdown condition is reached, but otherwise does not qualitatively change results, contrary to varying *β*, confirming that the large uncertainty in the initial conditions for the pandemic do not affect its dynamics. Thus it does not matter much to take for the initial condition, a single infected person in a specific group, or a specific amount of infected distributed over groups proportional to the population in each group, or a number proportional to the total amount of contacts each group makes. Furthermore, the infected category could be at different stages after their infection (Fig. S2D-F): due to the strong mixing induced by the contact matrix, the exact choice of initial conditions only affects the simulation time until lockdown conditions are reached, but otherwise has very little impact. Thus, after the lockdown time is properly calibrated in the simulations, the exact choice of the initial conditions has little influence. Because the pandemic was initially introduced from abroad, probably by persons in the age group 24-60 (group 2 and 3), we choose as initial condition for the number of infected per group 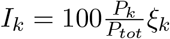, where ξ*_k_* = [0.5, 1, 0.5, 0, 0]. The total number of initially infected is ~ 53, comparable to what is used in [16].

#### Code availability

The codes are available upon request to the corresponding authors.

#### Author contributions

JR conceived the initial model, performed analysis and implemented the model in MATLAB. DH and JR refined and further developed the model and calibrated the model to data. AP and DH collected and analysed data. All authors contributed to writing of the manuscript.

#### Competing interests

The authors declare no competing interests.

## Acknowledgement

A. P. received funding from FRM (SPF201909009284), D. H. is supported by INSERM Plan Cancer and a Computational Neuroscience NIH-ANR grant. J. R. is supported by an ANR grant. We thanks Pr. Dan Longrois for some discussions about ICU’s organization.

---

**Table S1:** Parameter definitions,

| Parameter | Description |
| --- | --- |
| $n$ | Time in days |
| $m$ | Number of days after infection |
| $l$ | Number of days in current infection category. |
| $k$ | Group index. |
| $1 \leq J \leq J_{max}$ | Infection category including recovered category. |
| $1 \leq j \leq J_{max} - 1$ | Infection category without recovered category. |
| $a(m, l\vert k, j)$ | Probability to die. |
| $p(n, m, l\vert k, J_{old}, J_{new})$ | Transition probability to switch from category $J_{old}$ to $J_{new}$.<br>$\sum_{J_{new}} p(n, m, l\vert k, J_{old}, J_{new}) = 1$ (normalization) |
| $c(n\vert k, k')$ | Contact matrix at day $n$. |
| $\xi(m, l\vert k, j)$ | Matrix to identify infectious persons. |
| $P_k$ | Population of group $k$. |
| $\beta_k$ | Infectiousness of group $k$. |

**Table S2:** Population for the five regions. Source: https://www.statista.com/statistics/464032/distribution-population-age-group-france/

| $k$ | Group | Population (37M) |
| --- | --- | --- |
| 1 | 0-24 | 10.1M (30%) |
| 2 | 25-49 | 12.1M (33%) |
| 3 | 50-59 | 4.7M (13%) |
| 4 | 60-69 | 3.6M (10%) |
| 5 | 70+ | 5.5M (15%) |

**Table S3:**
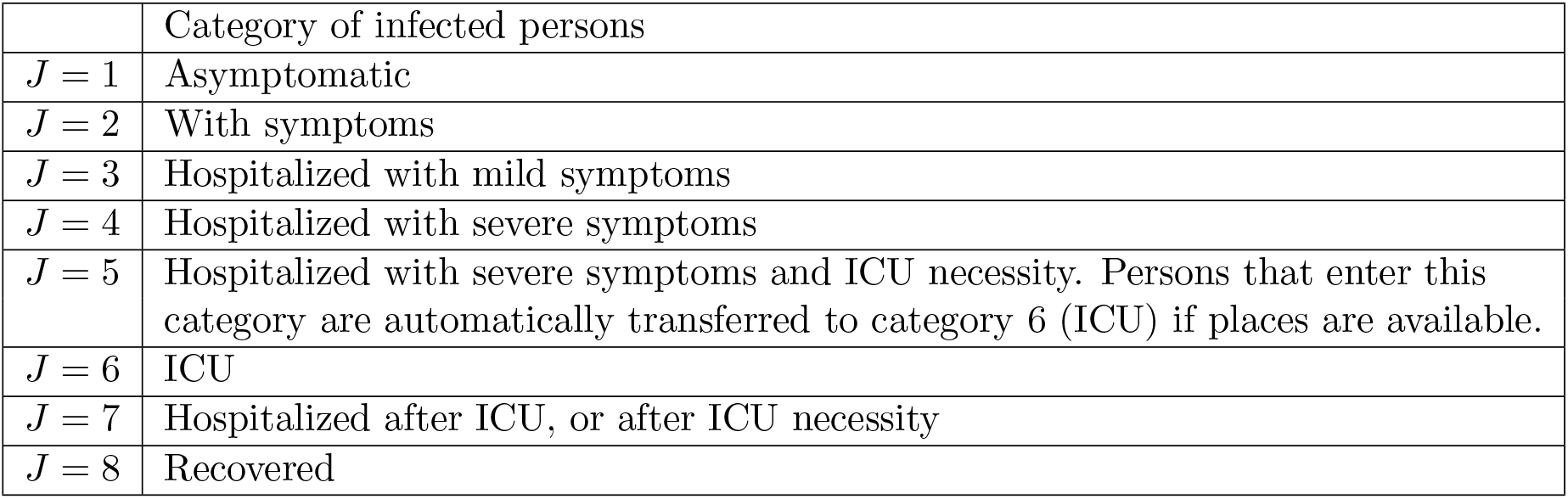
Infection categories that monitor the disease progression.

**Table S4:** Transition probabilities *p*(*n,m,l*|*k,J_old_, J_new_*). Only condition with non-zero values are specified. The probabilities to remain in the current category are 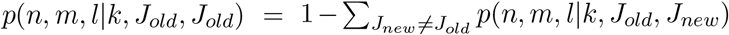. We assume a uniform transition probability per day during the specified transition period with duration *d_k_*. The daily probability is 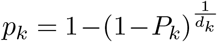. Hospitalized that switch to *j* = 5 are immediately transferred to *j* = 6 (ICU) unless ICU is full. We implemented the possibility to prioritise how liberating ICU places are distributed among the waiting persons in *j* = 5. Transitions to *J* = 7 and *J* = 8 occur with 95% probability during the specified period. The percentage that do not switch during this period change category the following day with probability *p_k_* = 1. This ensures that finally all persons leave ICU and the hospital. For group 5, the upper limit of the periods to recover from hospitalization (transition from *J* = 3, 4, 7 to *J* = 8) is gradually increased between 10-40 days after the lockdown daily by one day to comply with the published data for the daily number of hospitalisations for this group.

| $J_{old}$ | $J_{new}$ | Time | Transition period per group | Overall probability $P_k$ in % |
| --- | --- | --- | --- | --- |
| 1 | 2 | $l$ | ([6-10],[6-10],[6-10],[6-10],[6-10]) | (80, 80, 80, 90, 90) |
| 2 | 3 | $l$ | ([1-7],[1-7],[1-7],[1-7],[1-7]) | (0.16, 0.72, 2.7, 4.5, 30) |
| 3 | 4 | $l$ | ([1-3],[1-3],[1-3],[1-2],[1-2]) | (40, 70, 85, 75, 70) |
| 4 | 5 | $l$ | ([1],[1],[1],[1],[1]) | (20, 48, 45, 58, 45) |
| 5 | 6 | $l$ | | Automatic if ICU is available. |
| 5 | 7 | $l$ | ([8-10],[8-10],[8-10],[8-10],[8-10]) | (95, 95, 95, 95, 95) |
| 6 | 7 | $l$ | ([5-12],[5-12],[10-15],[10-18],[10-21]) | (95, 95, 95, 95, 95) |
| 1 | 8 | $l$ | ([12-18],[12-18],[12-18],[12-18],[12-18]) | (95, 95, 95, 95, 95) |
| 2 | 8 | $l$ | ([15-21],[15-21],[15-21],[15-21],[15-21]) | (95, 95, 95, 95, 95) |
| 3 | 8 | $l$ | ([4-7],[4-7],[5-10],[5-10],[5-10]) | (95, 95, 95, 95, 95) |
| 4 | 8 | $l$ | ([3-7],[3-7],[7-10],[10-14],[15-20]) | (95, 95, 95, 95, 95) |
| 7 | 8 | $l$ | ([3-8],[3-8],[7-14],[10-18],[14-21]) | (95, 95, 95, 95, 95) |

**Table S5:** Decease probabilities *a*(*m,l*|*k, j*). Persons die in the hospital in category *j* = 5 or *j* = 6. However, unless ICU is full, category *j* = 5 is empty. The per day probability is 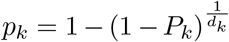.

| $j$ | Time | Period per group | Overall probability $P_k$ in % |
| --- | --- | --- | --- |
| 5 | $l$ | ([2-6],[2-6],[2-6],[2-6],[2-6]) | (80, 80, 80, 80, 90) |
| 6 | $l$ | ([2-8],[2-8],[2-8],[2-8],[2-5]) | (11, 13, 17, 30, 85) |

**Table S6:** Matrix ξ(*m,l*|*k, j*) to select persons that can infect others. We assume that only asymptomatic people infect others with uniform probability during 5-11 days after the infection. If persons show symptoms and switch to category *j* = 2, we assume that they put themselves in quarantine.

| $j$ | Time | Period per group | Value |
| --- | --- | --- | --- |
| 1 | $m$ | ([5-11],[5-11],[5-11],[5-11],[5-11]) | (1,1,1,1,1) |

## Data for different countries

Data plotted in figure S1 are taken from the repository managed by John Hopkins University https://github.com/CSSEGISandData/COVID-19. Four different countries have been considered: France, Italy, Germany and Czech Republic. Discontinuities in data are related to detection methods of each country. Curves are fitted using an exponential function, *f*(*t*) = α · *e^β·t^* + γ (dashed lines in the figure). Fits have been performed before confinement (17th March (France), 9th March (Italy), 22nd March (Germany) and 18th March (Czech Republic)) using Mathematica software, results are reported in the table.

**Table S7:** Fit exponents *β* before confinement

|  | France | Italy | Germany | Czech Republic |
| --- | --- | --- | --- | --- |
| Confirmed | $0.18 \pm 0.02$ | $0.19 \pm 0.01$ | $0.25 \pm 0.01$ | $0.26 \pm 0.01$ |
| Deaths | $0.18 \pm 0.03$ | $0.27 \pm 0.01$ | $0.36 \pm 0.02$ | xxx |

**Table S8:** Fitted exponents *β* in the first period after confinement. Fits after lockdown are performed using ten points (last point 10th April).

|  | France | Italy | Germany | Czech Republic |
| --- | --- | --- | --- | --- |
| Confirmed | $0.004 \pm 0.001$ | $0.003 \pm 0.001$ | $0.003 \pm 0.001$ | $0.003 \pm 0.001$ |
| Deaths | $0.005 \pm 0.001$ | $0.002 \pm 0.001$ | $0.09 \pm 0.01$ | $0.09 \pm 0.01$ |

## Hospitalization data

Data related to the hospitals after the confinement have been provided by French Government www.data.gouv.fr are reported in figure 1. In particular our analysis is based on the data in the following databases: *donnees-hospitalieres-covid19* and *donnees-hospitalieres-classe-age-covid19*. The first one contains the number of hospitalized people, currently in critical care (ICU), returned home and deaths divided for departments, the second the same data divided for regions and the relative age group distribution. In particular we focus our analysis on the most affected french regions: Île de France, Grand Est, Auvergne Rhône Alpes, Hauts-de-France and Provence-Alpes-Côte d’Azur. The age group distributions are available since the 18th of March (30 April is missing).

## Supplementary figures

**Figure S1:**
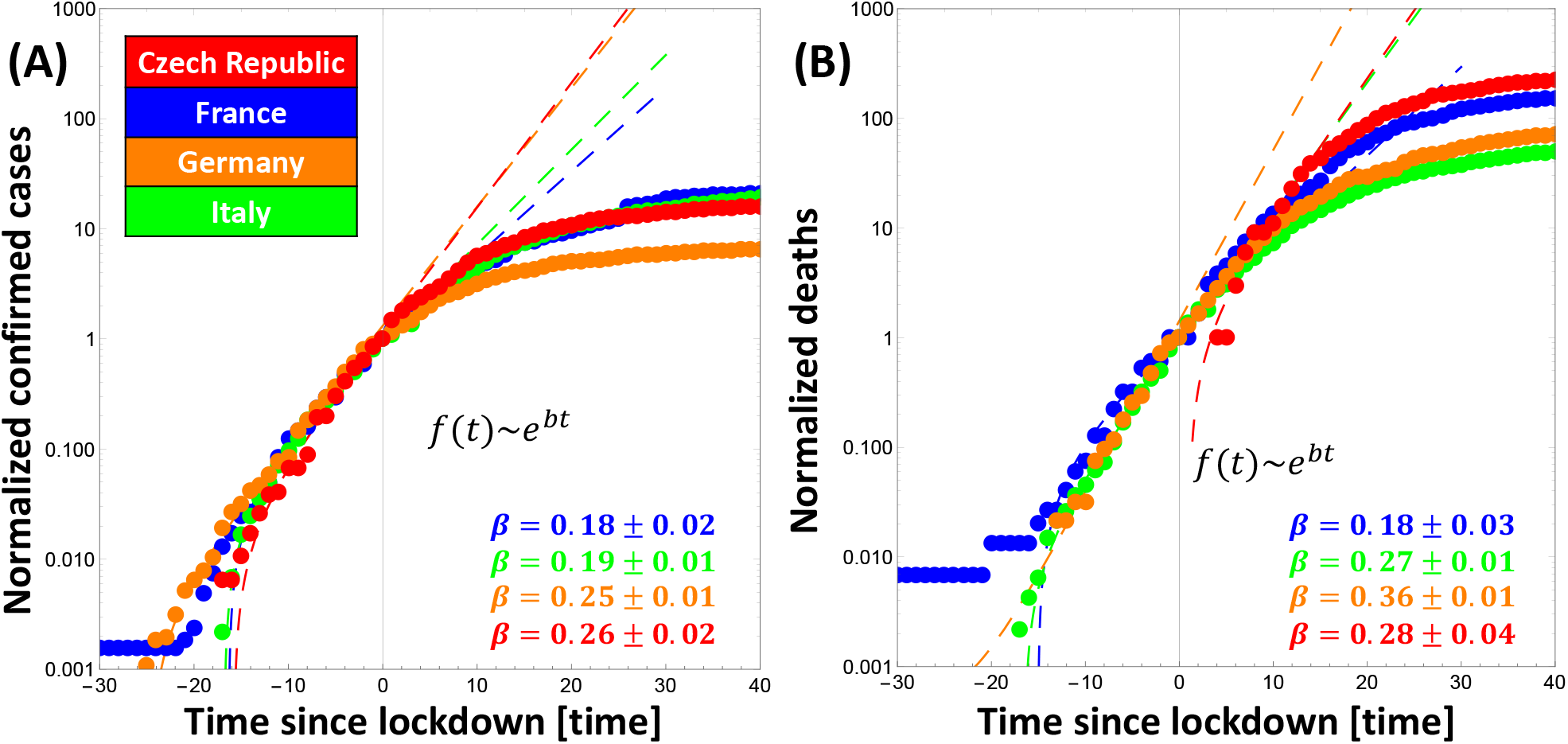
Exponential rates of confirmed cases (A) and deaths (B) before confinement for Czech Republic, France, Germany and Italy. Data are rescaled with values at the beginning of confinement. Since the number of deaths for Czech Republic on 18th March was zero, the curve in panel (B) is scaled to one.

**Figure S2:**
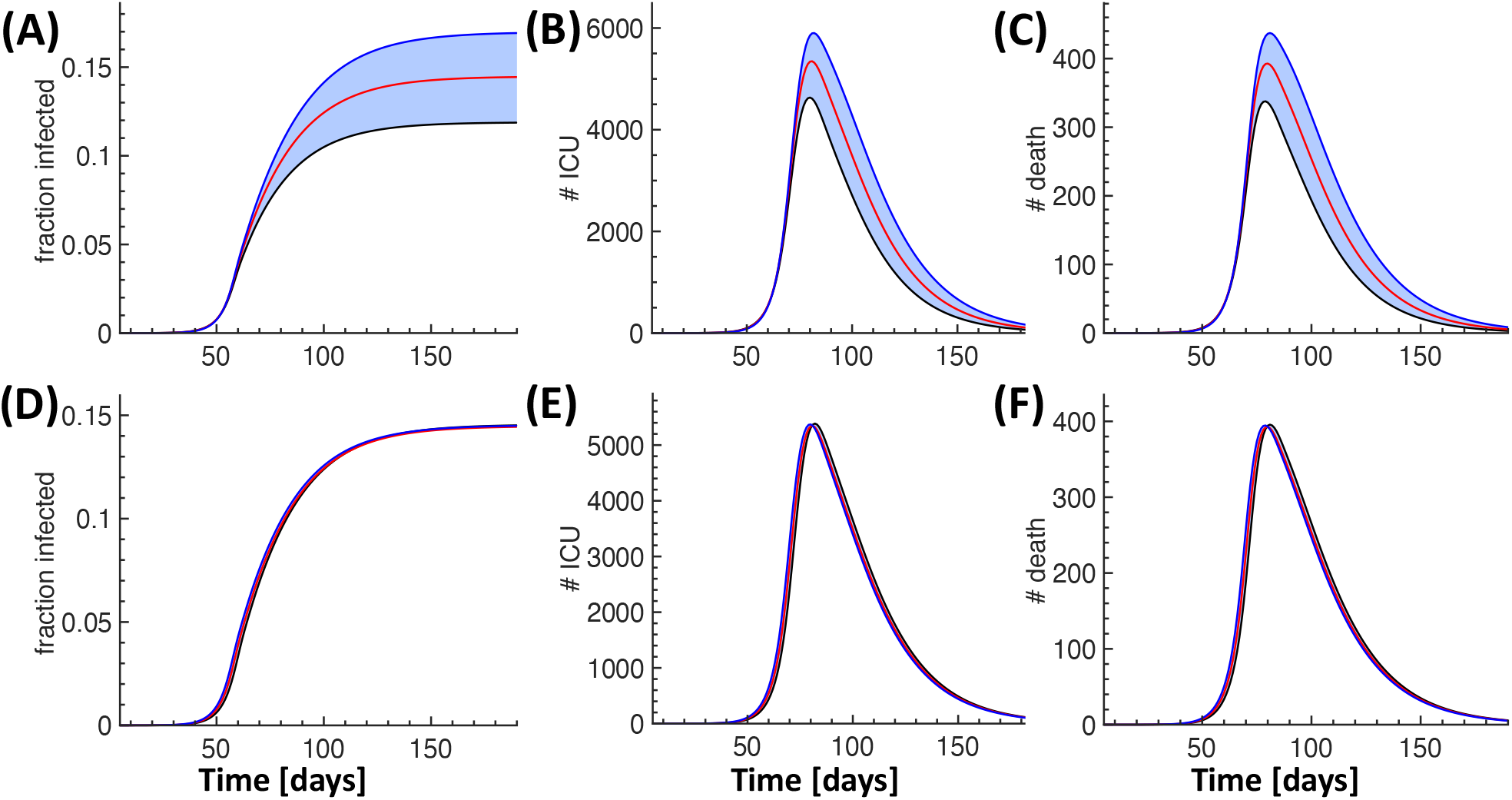
Sensitivity analysis for 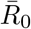 and initial conditions. Each panel shows three simulations for a scenario with lockdown and no deconfinement. For each simulation, the lockdown is imposed when the number of deceased reaches a fixed threshold. Simulation times are then aligned such that the lockdown occurs at day 61. (A-C) Consequences of varying the infectiousness 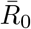 by ±5% for the fraction of infected (A), ICU occupancy (B) and deceased (C). (D-F) Consequences of varying the initial number of infected by ±20%.

**Figure S3:**
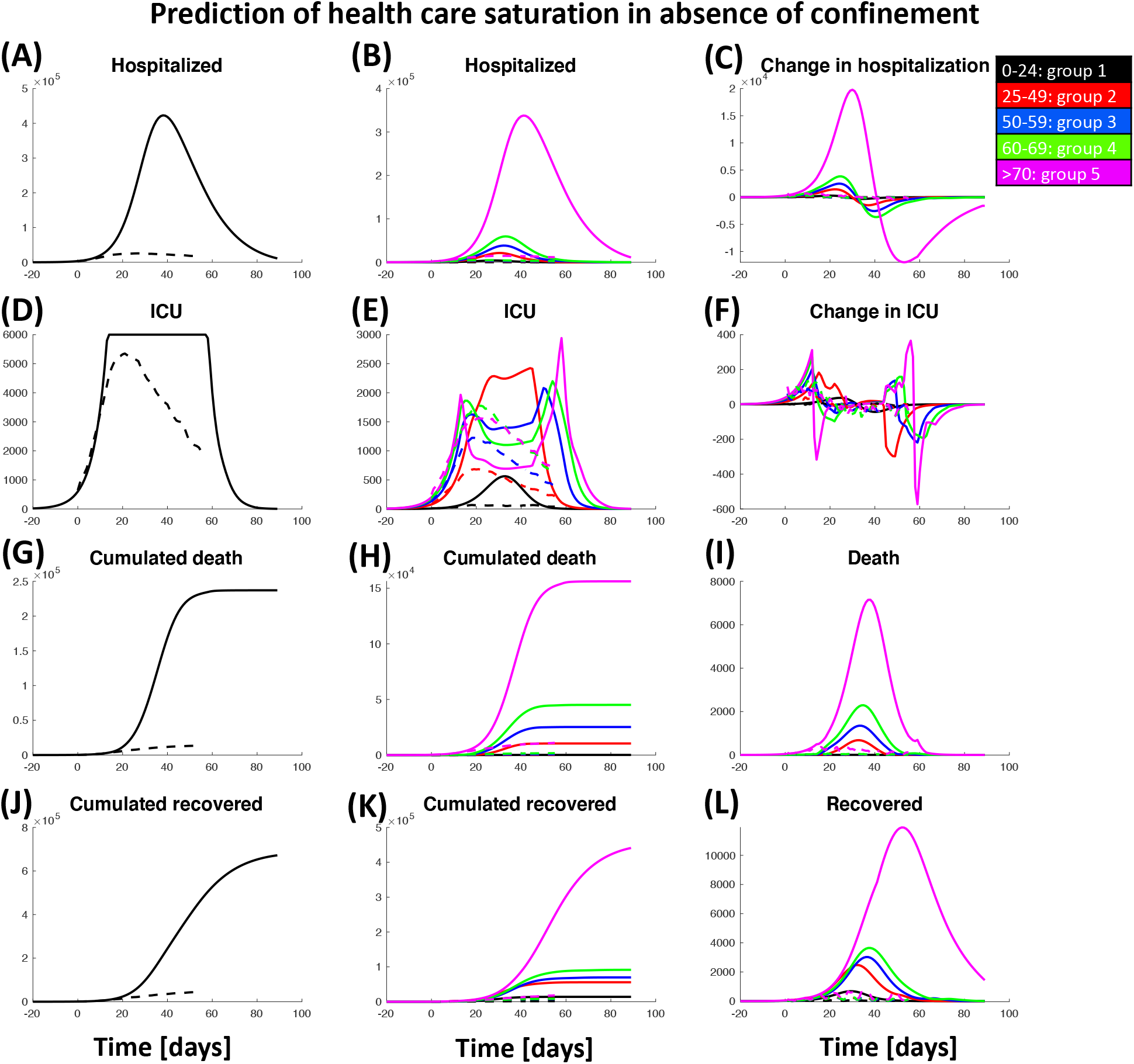
Prediction of pandemic spread and health care evolution in the absence of lockdown. Total number of hospitalized people (A), its age group distribution (B) and the daily variations (C); people in ICU (D), age group distribution in ICU (E) and the daily variations (F); cumulated deaths (G), age group distribution (H) and daily death (I); cumulative number of people recovered from hospitalization (J), age group distribution (K) and daily variations (L). Continuous lines show simulation results, dashed lines the actual data.

**Figure S4:**
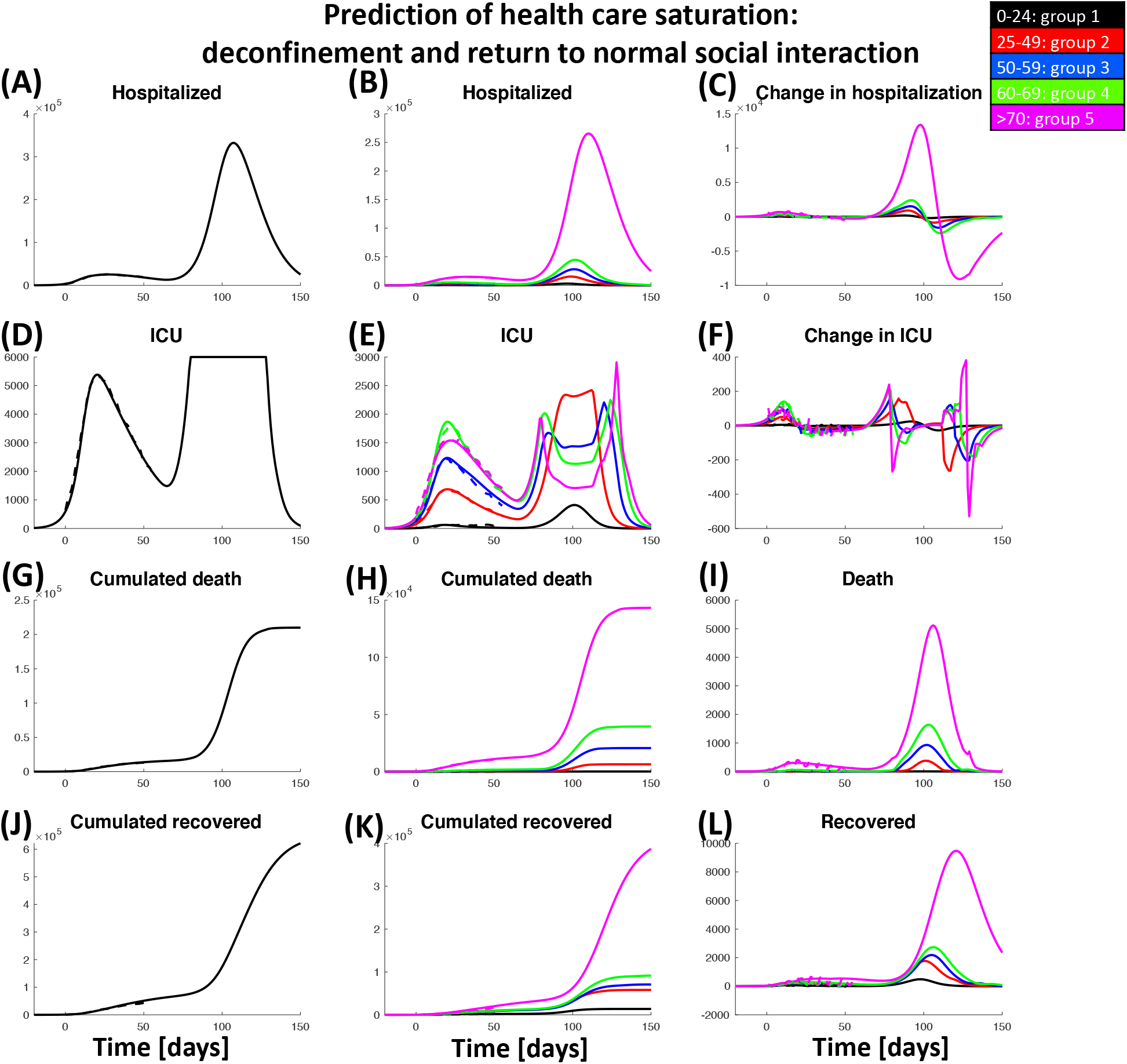
Prediction of pandemic spreading and health care saturation with full deconfinement and return to social interactions as before lockdown. Total number of hospitalized people (A), its age group distribution (B) and the daily variations (C); people in ICU (D), age group distribution in ICU (E) and the daily variations (F); cumulated deaths (G), age group distribution (H) and daily death (I); cumulative number of people recovered from hospitalization (J), age group distribution (K) and daily variations (L). Continuous lines show simulation results, dashed lines the actual data.

**Figure S5:**
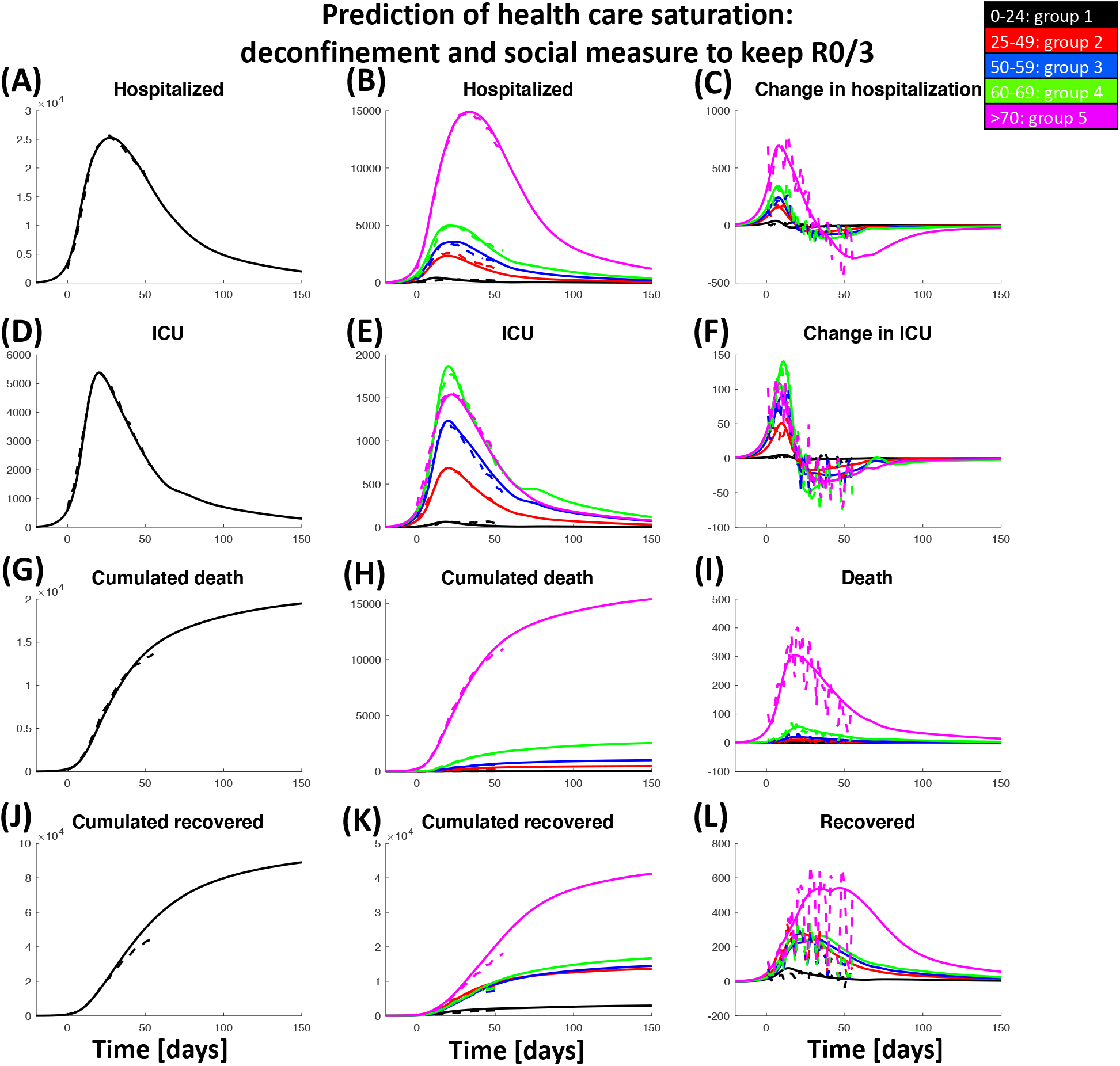
Predictions of health care evolution in a scenario with deconfinement and 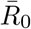 reduced by a factor of 3 compared to before lockdown. Total number of hospitalized people (A), its age group distribution (B) and the daily variations (C); people in ICU (D), age group distribution in ICU (E) and the daily variations (F); cumulated deaths (G), age group distribution (H) and daily death (I); cumulative number of people recovered from hospitalization (J), age group distribution (K) and daily variations (L). Continuous lines show simulation results, dashed lines the actual data.

**Figure S6:**
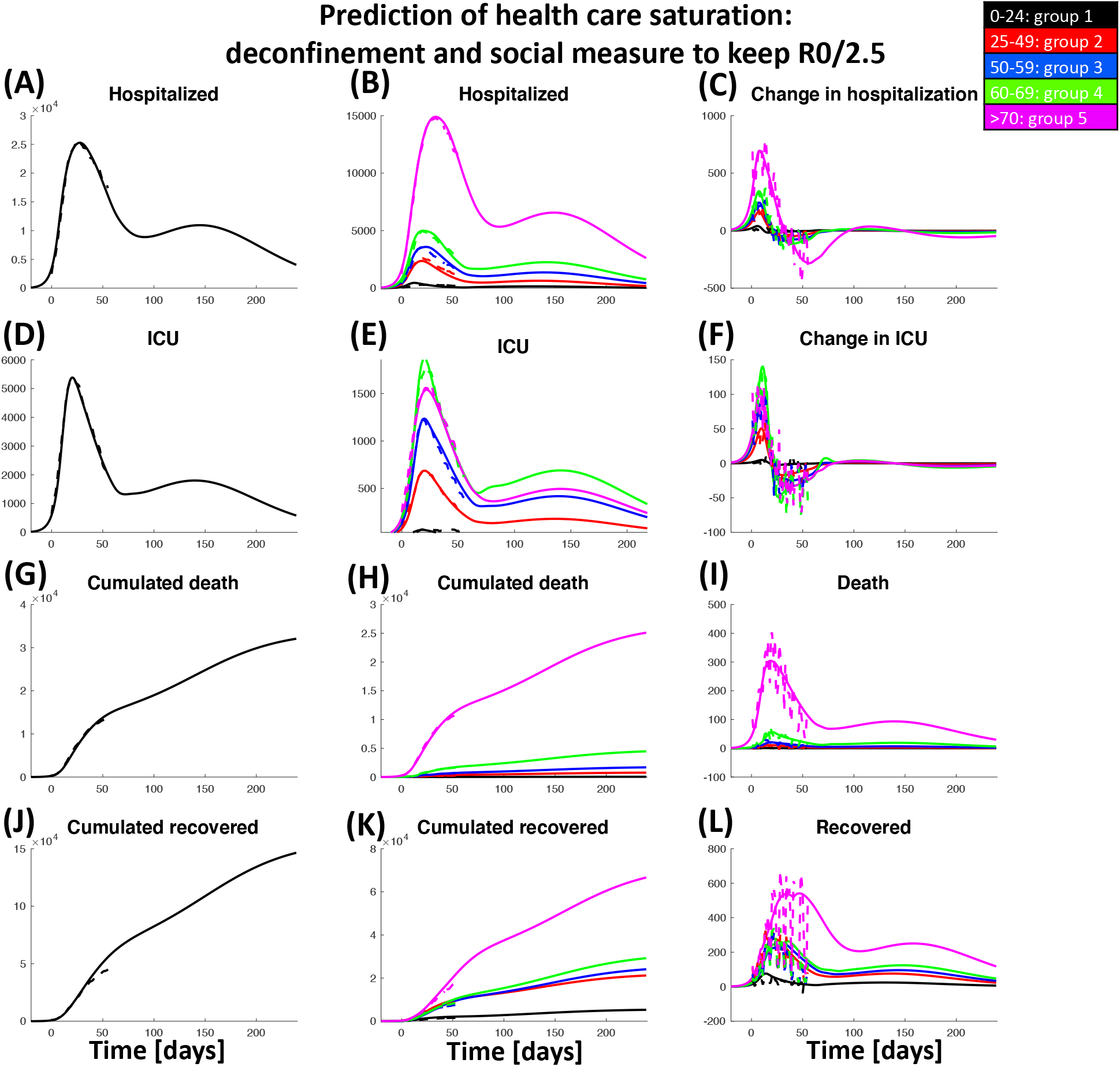
Predictions of health care evolution in a scenario with deconfinement and 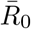 reduced by a factor of 2.5 compared to before lockdown. Total number of hospitalized people (A), its age group distribution (B) and the daily variations (C); people in ICU (D), age group distribution in ICU (E) and the daily variations (F); cumulated deaths (G), age group distribution (H) and daily death (I); cumulative number of people recovered from hospitalization (J), age group distribution (K) and daily variations (L). Continuous lines show simulation results, dashed lines the actual data.

## Notes

### Competing Interest Statement

The authors have declared no competing interest.

### Funding Statement

No funding

## References

[1] N. H. Leung, D. K. Chu, E. Y. Shiu, K.-H. Chan, J. J. McDevitt, B. J. Hau, H.-L. Yen, Y. Li, D. K. Ip, J. M. Peiris, et al., “Respiratory virus shedding in exhaled breath and efficacy of face masks,” Nature medicine, pp. 1-5, 2020.

[2] K. Leung, J. T. Wu, D. Liu, and G. M. Leung, “First-wave covid-19 transmissibility and severity in china outside hubei after control measures, and second-wave scenario planning: a modelling impact assessment,” The Lancet, 2020.

[3] Q. Li, X. Guan, P. Wu, X. Wang, L. Zhou, Y. Tong, R. Ren, K. S. Leung, E. H. Lau, J. Y. Wong, et al., “Early transmission dynamics in wuhan, china, of novel coronavirus-infected pneumonia,” New England Journal of Medicine, 2020.

[4] K. K. Cheng, T. H. Lam, and C. C. Leung, “Wearing face masks in the community during the covid-19 pandemic: altruism and solidarity,” The Lancet, 2020.

[5] R. Verity, L. C. Okell, I. Dorigatti, P. Winskill, C. Whittaker, N. Imai, G. Cuomo-Dannenburg, H. Thompson, P. G. T. Walker, H. Fu, A. Dighe, J. T. Griffin, M. Baguelin, S. Bhatia, A. Boonyasiri, A. Cori, Z. Cucunubá, R. FitzJohn, K. Gaythorpe, W. Green, A. Hamlet, W. Hinsley, D. Laydon, G. Nedjati-Gilani, S. Riley, S. [van Elsland], E. Volz, H. Wang, Y. Wang, X. Xi, C. A. Donnelly, A. C. Ghani, and N. M. Ferguson, “Estimates of the severity of coronavirus disease 2019: a model-based analysis,” The Lancet Infectious Diseases, 2020.

[6] J. T. Wu, K. Leung, and G. M. Leung, “Nowcasting and forecasting the potential domestic and international spread of the 2019-ncov outbreak originating in wuhan, china: a modelling study,” The Lancet, vol. 395, no. 10225, pp. 689-697, 2020.

[7] T. Jones, B. Mühlemann, T. Veith, M. Zuchowski, J. Hofmann, A. Stein, A. Edelmann, V. Max Corman, and C. Drosten, “An analysis of sars-cov-2 viral load by patient age,” Preprint, 2020.

[8] N. M. Ferguson, D. Laydon, G. Nedjati-Gilani, N. Imai, K. Ainslie, M. Baguelin, S. Bhatia, A. Boonyasiri, Z. Cucunubá, G. Cuomo-Dannenburg, A. Dighe, I. Dorigatti, H. Fu, K. Gaythorpe, W. Green, A. Hamlet, W. Hinsley, L. C Okell, S. van Elsland, H. Thompson, R. Verity, E. Volz, H. Wang, Y. Wang, P. G. Walker, C. Walters, P. Winskill, C. Whittaker, C. A. Donnelly, S. Riley, and A. C. Ghani, “Report 9: Impact of non-pharmaceutical interventions (npis) to reduce covid-19 mortality and healthcare demand,” Imperial College London, 2020.

[9] J. Dehning, J. Zierenberg, F. P. Spitzner, M. Wibral, J. P. Neto, M. Wilczek, and V. Priesemann, “Inferring change points in the covid-19 spreading reveals the effectiveness of interventions,” *medRxiv*, 2020.

[10] F. Balabdaoui and D. Mohr, “Age-stratified model of the covid-19 epidemic to analyze the impact of relaxing lockdown measures: nowcasting and forecasting for switzerland,” *medRxiv*, 2020.

[11] M. J. Keeling, E. Hill, E. Gorsich, B. Penman, G. Guyver-Fletcher, A. Holmes, T. Leng, H. McKimm, M. Tamborrino, L. Dyson, and M. Tildesley, “Predictions of covid-19 dynamics in the uk: short-term forecasting and analysis of potential exit strategies,” medRxiv, 2020.

[12] N. P. Jewell, J. A. Lewnard, and B. L. Jewell, “Caution warranted: Using the institute for health metrics and evaluation model for predicting the course of the covid-19 pandemic,” Annals of internal medicine, pp. M20–1565, 04 2020.

[13] I. for Health Metrics and Evaluation, “New covid-19 forecasts: Us hospitals could be overwhelmed in the second week of april by demand for icu beds, and us deaths could total 81,000 by july,” 2020.

[14] WorldHealthOrganization, “Population-based age-stratified seroepidemiological investigation protocol for covid-19 virus infection,” 2020.

[15] H. Streeck, B. Schulte, B. Kuemmerer, E. Richter, T. Hoeller, C. Fuhrmann, E. Bartok, R. Dolscheid, M. Berger, L. Wessendorf, M. Eschbach-Bludau, A. Kellings, A. Schwaiger, M. Coenen, P. Hoffmann, M. Noethen, A.-M. Eis-Huebinger, M. Exner, R. Schmithausen, M. Schmid, and B. Kuemmerer, “Infection fatality rate of sars-cov-2 infection in a german community with a super-spreading event,” *medRxiv*, 2020.

[16] H. Salje, C. Tran Kiem, N. Lefrancq, N. Courtejoie, P. Bosetti, J. Paireau, A. Andronico, N. Hoze, J. Richet, C.-L. Dubost, Y. Le Strat, J. Lessler, D. L. Bruhl, A. Fontanet, L. Opatowski, P.-Y. Boëlle, and S. Cauchemez, “Estimating the burden of sars-cov-2 in france,” *medRxiv*, 2020.

[17] “https://geodes.santepubliquefrance.fr.”

[18] “https://www.data.gouv.fr/fr/datasets/donnees-hospitalieres-relatives-a-lepidemie-de-covid-19.”

[19] G. Béraud, S. Kazmercziak, P. Beutels, D. Levy-Bruhl, X. Lenne, N. Mielcarek, Y. Yazdanpanah, P.-Y. Boelle, N. Hens, and B. Dervaux, “The french connection: The first large population-based contact survey in france relevant for the spread of infectious diseases,” PLOS ONE, vol. 10, no. 7, pp. 1-22, 2015.

[20] L. Di Domenico, G. Pullano, C. E. Sabbatini, Boëlle, Pierre-Yves, and V. Colizza, “Report 9, expected impact of lockdown in île-de-france and possible exit strategies,” 2020.

[21] P. Anfinrud, V. Stadnytskyi, C. E. Bax, and A. Bax, “Visualizing speech-generated oral fluid droplets with laser light scattering,” New England Journal of Medicine, 2020.

[22] D. K. Milton, M. P. Fabian, B. J. Cowling, M. L. Grantham, and J. J. McDevitt, “Influenza virus aerosols in human exhaled breath: Particle size, culturability, and effect of surgical masks,” PLOS Pathogens, vol. 9, no. 3, pp. 1-7, 2013.

